# Cardiovascular disease and Osteoporosis : A Mendelian randomization study

**DOI:** 10.1101/2024.01.09.24300808

**Authors:** Zhao Wang, Shuyi Zhang, Hongyang Gong, Guoxu Zhao, Hong Xiao, Shuai Yuan, Wenhao Wu, Pai Xu, Yihong Chen, Mei Yang, Chan Kang

**Affiliations:** Department of Orthopedic Surgery, Chungnam National University School of Medicine, Daejeon, South Korea; Department of Orthopedics, Fuzhou second Hospital, Fuzhou, Fujian, China; Department of Physiology, College of Medicine, Chosun University, Gwangju, South Korea; School of medical imaging, Mudanjiang Medical College, Heilongjiang Province, China; Department of Orthopedic, Southwest Hospital Jiangbei Area (The 958th hospital of Chinese People’s Liberation Arm y), Jianxindong Road, Jiang Bei District, Chongqing 400020, China; Department of Hepatobiliary Pancreatic Surgery, Changhai Hospital, Second Military Medical University (Naval Medic al University), Shanghai, China; Intervention Center Ward Unit 2,Tangdu Hospital, Air Force Medical University, Xi’an, China; Department of Clinical Medicine, Xiamen Medical College, Xiamen, China; School of Health Science, Mae Fah Luang University, Thailand

**Author notes:** These authors contributed equally to this work and share first authorship (Zhao Wang, Shuyi Zhang, Hongyang Gong, Guoxu Zhao).

## Abstract

I.

**Background:** Cardiovascular disease (CVD) may have some association with osteoporosis (OP). This Mendelian randomization (MR) investigation aimed to explore the potential causal linkage between CVD and OP.

**Methods:** Utilizing genome-wide association study data from individuals of European descent, we pinpointed Single Nucleotide Polymorphisms (SNPs) relevant to CVD, including those for coronary heart disease (CHD) with 64,762 cases and 22,233 controls, heart failure (HF) comprising 47,309 cases against 930,014 controls, and stroke with a case-control tally of 3,611 to 18,084, to serve as the instrumental variables. Later, we searched for total body bone mineral density (BMD) statistics which were used as phenotypes for OP(sample size = 56,284). In this paper, the traditional inverse variance weighting (IVW) method, the weighted median estimation method, and the MR-Egger method are used to estimate different results. The MR-Egger intercept test, outlier (Mr-PRESSO) test and Cochran-Q statistic are used to detect potential directional pleiotropy and heterogeneity, while we also draw the scatter plot, funnel plot and forest plot. Additionally, a reverse-direction MR analysis was performed to explore the potential for reverse causation.

**Results:** The IVW analysis showed that CHD could significantly impact total body BMD levels, and every higher standard deviation in the risk of CHD decreased the average total body BMD by 0.0459 units in the IVW analysis(Beta = -0.0459; 95%CI = -0.0815 –-0.0104, P = 0.0113). Reverse MR analysis showed no significant correlation of the change of total body BMD on the prevalence effect of CHD. No particular relationship exists between HF and total body BMD. There was no significant effect between the changes in total body BMD induced by stroke. Reverse MR analysis revealed no significant correlation between alterations in total body BMD on stroke.

**Conclusion:** Our analysis points to a substantial causative link between CHD and the vulnerability to OP, potentially paving the way for innovative approaches in treating and preventing OP.

## II. Introduction

### 1. Cardiovascular diseases

Within the landscape of global disease burden, the impact of cardiovascular disease (CVD) is becoming increasingly prominent, not only due to its direct threat to human health but also because of their profound socio-economic repercussions. CVD is the leading cause of death worldwide, with nearly 20 million people succumbing to it in 2017, a figure that far exceeds the global toll of COVID-19^1^. Accompanying demographic shifts toward an aging population and changes in lifestyle, the prevalence of CVD is on the rise, presenting unprecedented challenges for individuals, families, and society at large.

### 2. Osteoporosis

Osteoporosis (OP) is a chronic metabolic bone disease characterized by reduced bone mass, microarchitectural deterioration of bone tissue, reduced bone tension and strength, and increased risk of fragility fractures^2^. The diagnosis of OP is based on bone mineral density (BMD), usually measured by dual-energy X-ray absorptiometry^3^. According to World Health Organization guidelines, OP is defined as a BMD value less than 2.5 standard deviations below the mean (T-score) of a young, healthy population matched by sex and ethnicity^4^. The global prevalence of OP is 18.3%, with large differences among different ethnic groups and regions^5^. Among the consequences of OP, fractures are one of the most serious and result in a huge economic burden^6^. Therefore, there is a need to explore potential causal risk factors for OP.

### 3. CVD and OP

The relationship between CVD and OP has become a hot topic of research in the medical field. However, the exact nature of the relationship is still controversial. Many studies have explored the potential relationship between CVD and BMD or fracture risk, but the results are not consistent.

Many large-scale epidemiological studies have found an association between coronary heart disease (CHD) and OP. For example, Marcovitz PA et al. have shown that low BMD can predict coronary artery disease^7^. For another example, the study of DANIELLE L et al. found that patients with CHD were more likely to have problems in BMD^8^. The study based on the third National Health and Nutrition Examination Survey, and patients at higher risk of CHD also had a generally higher risk of lower BMD. In the research by Tekin and colleagues, it was observed that among female subjects, there was no correlation identified between reduced BMD and CHD, as assessed by angiographic evaluation in those undergoing coronary angiography^9^. The latest longitudinal cohort analysis from Zhu and colleagues indicates that overall BMD might independently predict the likelihood of stroke incidents, particularly among male subjects^10^. However, another study incorporating both prospective research and a meta-analysis has drawn conflicting conclusions^11^.

### 4. Mendelian randomization

Epidemiology is the scientific study of the distribution of diseases and health outcomes within specific populations^12^. The term “classical epidemiology” is used to denote epidemiologic research that does not involve genetics, as opposed to genetic epidemiology. Randomized controlled trials (RCTs) are often considered the gold standard for testing scientific hypotheses in clinical research. While RCTs are theoretically the optimal method for establishing clear causal relationships between specific exposures and outcomes, they have inherent limitations^13^. RCTs are costly and time-consuming, especially when the outcomes of interest are rare or require long-term follow-up. Additionally, it may not always be feasible to intervene solely on the exposure of interest. Moreover, due to practical or ethical reasons, random allocation for certain exposures may not be possible. Observational studies are more readily conducted and often test scientific hypotheses by comparing outcome distributions at varying levels of exposure without intervention. However, differences in outcome distributions between two cohorts can sometimes be incorrectly interpreted as causal effects of exposure. Such conclusions may confound correlation with causation.

Mendelian randomization (MR) design can use observational data to conduct causal association studies, providing an effective way for epidemiological causal inference^14^. In 1986, Katan proposed the concept of MR^15^. The fundamental idea is to use genetic variants, such as single nucleotide polymorphisms (SNPs), that are strongly correlated with the exposure as instrumental variables (IVs) for modeling to estimate the causal relationship between exposure and outcome. Mendel’s law of inheritance describes the random allocation of parental alleles to offspring, rendering gamete formation akin to a “natural” randomized controlled trial that mitigates confounding from factors like living environment, social influences, and acquired behaviors. This random allocation, completed before birth, ensures the proper causal sequence. Hence, the MR method plays a pivotal role in causal inference in epidemiology.

Currently, the MR method is rapidly advancing, classified into two main types based on data characteristics: single-sample MR and two-sample MR. Single-sample MR requires both exposure and outcome to originate from the same dataset, and its establishment demands significant resources, leading to high costs. To overcome this limitation, the two-sample MR was introduced^16^, allowing the exposure and outcome to come from two distinct datasets, and enabling the evaluation of the causal effect of exposure on outcome in two non-overlapping studies. Two-sample MR analysis utilizes existing Genome-Wide Association Studies (GWAS) summary data. Compared to individual data, summary data is more accessible, without ethical constraints. Publicly available summary data also enhances the utility of bioinformatics, thereby reducing research costs.

GWAS analyzes the correlation between complex traits and millions of molecular markers at whole genome level: specifically, SNPs. By contrasting, it identifies genetic variations influencing these complex traits^17^. The rapid advancement of GWAS has made two-sample MR an efficient and economical method to explore the causal relationships between health risk factors and disease outcomes.

Thus, this study uses publicly published GWAS summary data, adopts the two-sample MR framework to investigate the causal relationship between CVD and OP. Additionally, by employing a reverse MR analysis, we further validate the study findings, providing fresh insights for the etiological research of OP.

## III. Methods

### 1. Study overview

In our MR examination, total body BMD was chosen as the focal point of study. To encapsulate CVD, we specifically included the most prevalent conditions: CHD, heart failure (HF), and stroke. For the direct MR analysis, SNPs with a high correlation to CHD, HF, and stroke were employed as the IVs. Conversely, in the indirect MR analysis, SNPs with a significant association to total body BMD were utilized. Figure 1 illustrates the schematic of the research approach.

**FIGURE 1.**
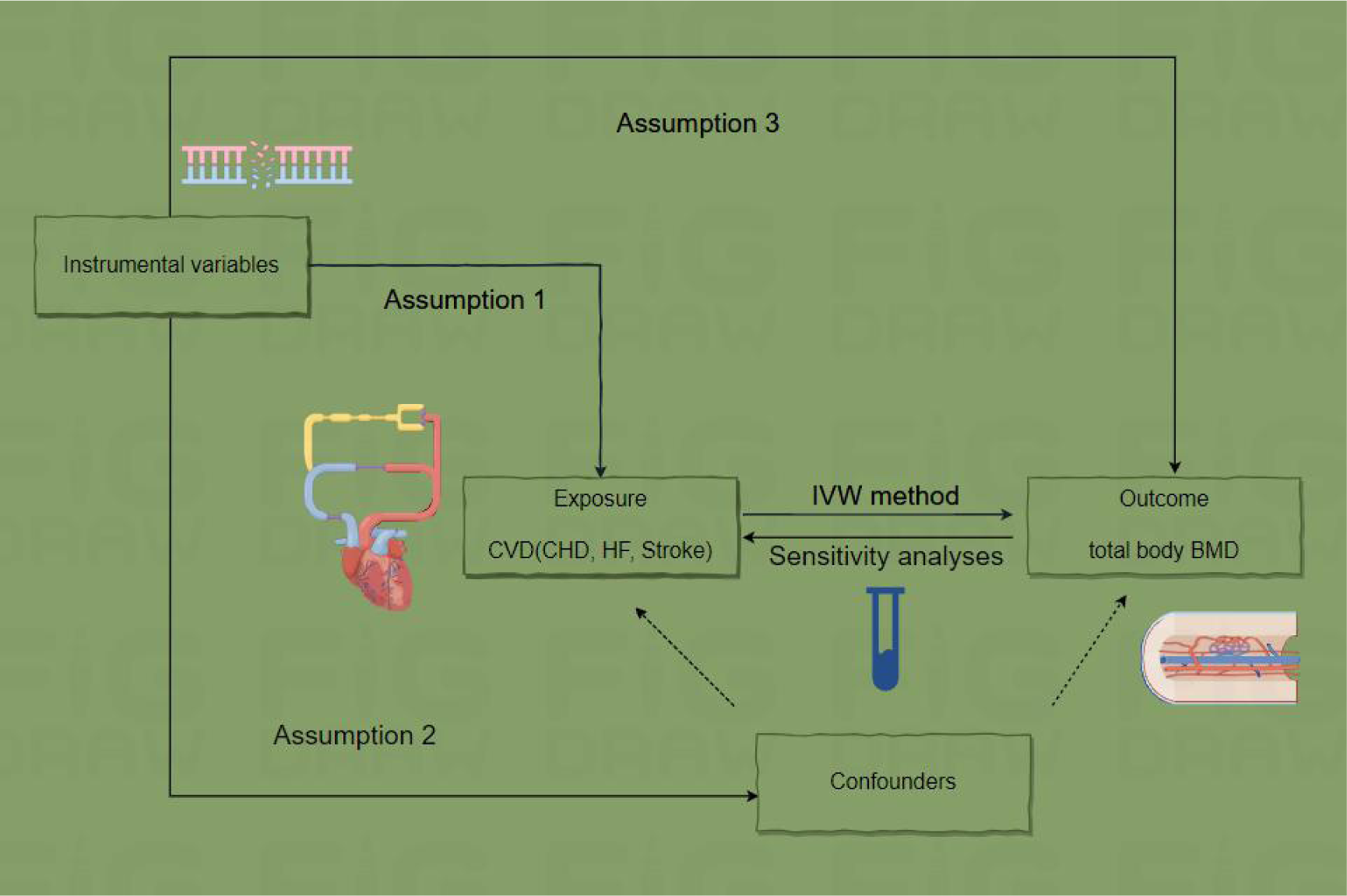
Conceptual Framework of the Study Design. ‘SNP’ denotes Single Nucleotide Polymorphism, ‘CHD’ stands for coronary heart disease, ‘HF’ represents heart failure, ‘BMD’ refers to bone mineral density, and ‘IVW’ indicates inverse-variance weighted. The diagram is founded on three critical assumptions: (1) Instrumental variables exhibit a strong correlation with the intended risk factor; (2) Instrumental variables maintain independence from confounding factors; (3) Instrumental variables influence the outcome exclusively via the risk factor, not through secondary pathways. Sensitivity checks, including MR-PRESSO and the leave-one-out analysis, are illustrated as well.

### 2. Data Source and Definition

#### A. GWAS summary statistics for BMD

Data for BMD were obtained from the GWAS database (https://gwas.mrcieu.ac.uk) of the Integrated Epidemiology Unit (IEC). We selected BMD (sample size = 56,284). The raw data can be found in the Medina-Gomez C study^18^. All races selected from the above data are of European ancestry.

#### B. GWAS summary statistics for CVD (CHD, HF, Stroke)

We selected the data on BMD also based on the large GWAS database. Data for CHD are relevant from Schunkert H studies exploring genetic variation in 86,995 Europeans (64,762 cases, 22,233 controls)^19^. Data for HF can be found in the Shah S article^20^ (47309 cases, 930014 controls). Stroke was chosen as a variable, with 3,611 instances and 18,084 control entries. Malik R’s study ^21^ is the source for the original dataset.

### 3. Instrumental Genetic Variables

We established a consistent selection threshold for the IVs to adhere to the first principle of MR. First, P <1×10^-5^ was selected as inclusion condition to obtain statistically significant association analysis results. Second, we performed a linkage disequilibrium analysis of the SNPs corresponding to each instrumental variable (r^2^ = 0.001, kb = 10000) to ensure that the SNP was independent. To rigorously assess the secondary and tertiary presuppositions of MR, we mitigated the impact of acknowledged confounders on causal inference. Moreover, SNPs linked to CVD were meticulously excised via a phenotype scanning tool available at [http://www.phenoscanner.medschl.cam.ac.uk].

Identified confounders for CVD include factors such as lipids, body mass index (BMI), smoking, alcohol consumption, physical activity, sleep duration, educational level, and hypertension^22^.

Potential confounders associated with BMD include alcoholism, body mass index(BMI), glucocorticoid overdose, hypogonadism, parathyroidism, hyperthyroidism, gastro-intestinal diseases, hypercalciuria, fasting insulin levels, type 2 diabetes, fasting glucose levels, HDL cholesterol levels, depression, sex hormones, lifestyle factors^23^.

After screening, we found and excluded some of these IV about BMI in the sensitivity model, and reanalyzed the MR estimates to obtain a more direct causal association.

For each SNP, the F-statistic was derived employing the formula:

F = [R2combined / (1 - R2combined)] * [(N - K - 1) / K]

Here, R2 is determined using the equation:

R^2^ = [beta. exposure^2^] / [se. exposure^2^ * N + beta. exposure^2^]

In the above formula, K = number variants comprising instrument, and N = GWAS sample size, beta. exposure = effect of SNP on exposure, se. exposure = standard error for the SNP’s exposure effect. The F statistic evaluates the strength of the tool, and the SNP with F < 10 is considered weak and removed^24^.

### 4. Quantitative Analysis

We investigated the potential causal influence of CVD (CHD, HF, Stroke) on total body BMD, followed by a reverse MR examination to discern the influence of total body BMD on CVD (CHD, HF, Stroke), thereby constituting a bidirectional MR investigation.

The inverse-variance weighted (IVW) method was employed as the principal approach for our analysis ^25^. The IVW method is an ideal estimate and has a strong ability to detect causality. Two additional MR models were also used the weighted median and the MR-Egger method^26, 27^. They can tolerate the presence of horizontal pleiotropy but have lower statistical power than IVW. The presence of horizontal pleiotropy was estimated by assessing the MR-Egger regression intercept^28^. Furthermore, the Cochran’s Q test was used to assess the heterogeneity between the SNPs in the IVW estimates (P > 0.05 indicates no significant heterogeneity). When there is no heterogeneity, we use a fixed effect model, and if heterogeneity is found, we use a random effect model^29^. If heterogeneity is detected in the MR-PRESSO test ^30^, then we will remove the outlier variant (P < 0.05 in the MR-PRESSO test) and perform the MR analysis again. For sensitivity analyses, we used the leave-one-out test ^31^ and, after removal, revisited the MR analysis if any variables influenced the causal effect estimates in order to assess potential violations of these assumptions. The influential SNPs were evaluated from two plots (forest, and scatter). All statistical data analyses were performed using R software, version 4.1.3, using the R package for “TwoSampleMR” and “MR-PRESSO”^30, 32^ and considering for difference with P < 0.05. Ethical approvals and informed consents for this study can be found in the respective original publications associated with the utilized public databases.

## IV. Results

### 1. Influence of CVD (CHD, HF, Stroke) on total body BMD: A Causal Assessment

Confounding genes removed are shown in Table 1, 2, 3. Details used for the SNPs related to CVD (CHD, HF, Stroke) and the SNPs associated with total body BMD are listed in Table 4, 5, 6.

**Table 1.** The removed SNPs containing known confounding factors of CHD on total body BMD.

| SNP | Chr | Pos | EA | OA | CHD (Exposure) |  |  | Total body BMD (Outcome) |  |  |
| --- | --- | --- | --- | --- | --- | --- | --- | --- | --- | --- |
|  |  |  |  |  | Beta | SE | P-value | Beta | SE | P-value |
| rs1333045 | 9 | 22119195 | C | T | 0.226084 | 0.0058 | 0.8407 | 0.0012 | 0.0058 | 0.8407 |
| rs7651039 | 3 | 15648004 | C | T | 0.142119 | 0.0058 | 0.5386 | -0.0036 | 0.0058 | 0.5386 |
| rs964184 | 11 | 116648917 | C | G | -0.12596 | 0.0082 | 0.1916 | 0.0107 | 0.0082 | 0.1916 |

Abbreviations: SNP,single-nucleotide polymorphism; Chr, chromosome; Pos, position; EA, effect allele; OA, other allele; SE, standard error; BMD, bone mineral density; CHD, coronary heart disease.

**Table 2.** The removed SNP containing known confounding factors of HF on total body BMD.

| SNP | Chr | Pos | EA | OA | HF (Exposure) |  |  | Total body BMD (Outcome) |  |  |
| --- | --- | --- | --- | --- | --- | --- | --- | --- | --- | --- |
|  |  |  |  |  | Beta | SE | P-value | Beta | SE | P-value |
| rs56094641 | 16 | 53806453 | G | A | 0.0454 | 0.008 | 1.20801E-08 | -0.012 | 0.0059 | 0.0401698 |

Abbreviations: SNP,single-nucleotide polymorphism; Chr, chromosome; Pos, position; EA, effect allele; OA, other allele; SE, standard error; BMD, bone mineral density; HF, heart failure.

**Table 3.** The removed SNP containing known confounding factors of stroke on total body BMD.

| SNP | Chr | Pos | EA | OA | Stroke (Exposure) |  |  | Total body BMD (Outcome) |  |  |
| --- | --- | --- | --- | --- | --- | --- | --- | --- | --- | --- |
|  |  |  |  |  | Beta | SE | P-value | Beta | SE | P-value |
| rs3184504 | 12 | 111884608 | C | T | -0.0779 | 0.0101 | 1.22914E-14 | 0.0193 | 0.0058 | 0.000845493 |

Abbreviations: SNP,single-nucleotide polymorphism; Chr, chromosome; Pos, position; EA, effect allele; OA, other allele; SE, standard error; BMD, bone mineral density.

**Table 4.** Association of the SNPs used as candidate genetic instruments from the GWAS for Mendelian randomization analyses of CHD and risk of total body BMD.

| SNP | Chr | Pos | EA | OA | CHD (Exposure) |  |  | Total body BMD (Outcome) |  |  |
| --- | --- | --- | --- | --- | --- | --- | --- | --- | --- | --- |
|  |  |  |  |  | Beta | SE | P-value | Beta | SE | P-value |
| rs10455872 | 6 | 161010118 | G | A | 0.27787 | 0.038112 | 3.08035E-13 | -0.023 | 0.0127 | 0.0711803 |
| rs1122608 | 19 | 11163601 | T | G | -0.127428 | 0.0208429 | 9.72994E-10 | 0.0004 | 0.0067 | 0.9498 |
| rs11556924 | 7 | 129663496 | T | C | -0.0905122 | 0.0151329 | 2.21998E-09 | 0.0133 | 0.0062 | 0.0328897 |
| rs12190287 | 6 | 134214525 | G | C | -0.103209 | 0.0156807 | 4.63981E-11 | 0.0023 | 0.0063 | 0.713199 |
| rs1333045 | 9 | 22119195 | C | T | 0.226084 | 0.019183 | 4.6302E-32 | 0.0012 | 0.0058 | 0.8407 |
| rs17114036 | 1 | 56962821 | G | A | -0.144965 | 0.0255686 | 1.43001E-08 | 0.004 | 0.0096 | 0.6773 |
| rs2219939 | 15 | 79029723 | A | G | -0.0990217 | 0.016288 | 1.21001E-09 | -0.007 | 0.007 | 0.3202 |
| rs2306374 | 3 | 138119952 | C | T | 0.108439 | 0.019636 | 3.34003E-08 | -0.0161 | 0.0078 | 0.0397201 |
| rs2351524 | 2 | 203880992 | C | T | -0.13869 | 0.0206229 | 1.75995E-11 | -0.0009 | 0.0088 | 0.9145 |
| rs4714955 | 6 | 12903435 | T | C | -0.0997862 | 0.0145058 | 6.02976E-12 | 0.0006 | 0.0063 | 0.9245 |
| rs599839 | 1 | 109822166 | A | G | 0.106715 | 0.0169259 | 2.89001E-10 | -0.0142 | 0.0067 | 0.0337598 |
| rs7651039 | 3 | 15648004 | C | T | 0.142119 | 0.0252631 | 1.84999E-08 | -0.0036 | 0.0058 | 0.5386 |
| rs9351814 | 6 | 72193707 | C | A | -0.0795946 | 0.0141883 | 2.01999E-08 | 0.0005 | 0.0058 | 0.9358 |
| rs964184 | 11 | 116648917 | C | G | -0.12596 | 0.0204991 | 8.01992E-10 | 0.0107 | 0.0082 | 0.1916 |
| rs9982601 | 21 | 35599128 | T | C | 0.163991 | 0.0262564 | 4.21998E-10 | -0.007 | 0.0085 | 0.4115 |

Abbreviations: SNP, single-nucleotide polymorphism; Chr, chromosome; Pos, position; EA, effect allele; OA, other allele; SE, standard error; BMD, bone mineral density; CHD, coronary heart disease.

**Table 5.** Association of the SNPs used as candidate genetic instruments from the GWAS for Mendelian randomization analyses of HF and risk of total body BMD.

| SNP | Chr | Pos | EA | OA | HF (Exposure) |  |  | Total body BMD (Outcome) |  |  |
| --- | --- | --- | --- | --- | --- | --- | --- | --- | --- | --- |
|  |  |  |  |  | Beta | SE | P-value | Beta | SE | P-value |
| rs11745324 | 5 | 137012171 | A | G | -0.0528 | 0.0095 | 2.34498E-08 | 0.0064 | 0.0069 | 0.3533 |
| rs1510226 | 6 | 160816409 | C | T | 0.162 | 0.0285 | 1.26599E-08 | 0.0079 | 0.0211 | 0.709999 |
| rs1556516 | 9 | 22100176 | C | G | 0.0622 | 0.0078 | 1.56892E-15 | 0.001 | 0.0057 | 0.8585 |
| rs17042102 | 4 | 111668626 | A | G | 0.1103 | 0.0121 | 5.70558E-20 | -0.0056 | 0.009 | 0.530401 |
| rs17617337 | 10 | 121426884 | T | C | -0.0561 | 0.0095 | 3.65401E-09 | -0.0019 | 0.007 | 0.789 |
| rs4746140 | 10 | 75417249 | C | G | -0.0666 | 0.0109 | 1.104E-09 | 0.006 | 0.0076 | 0.4308 |
| rs55730499 | 6 | 161005610 | T | C | 0.1058 | 0.0157 | 1.83021E-11 | -0.0212 | 0.0126 | 0.0933405 |

Abbreviations: SNP, single-nucleotide polymorphism; Chr, chromosome; Pos, position; EA, effect allele; OA, other allele; SE, standard error; BMD, bone mineral density; HF, heart failure.

**Table 6.**
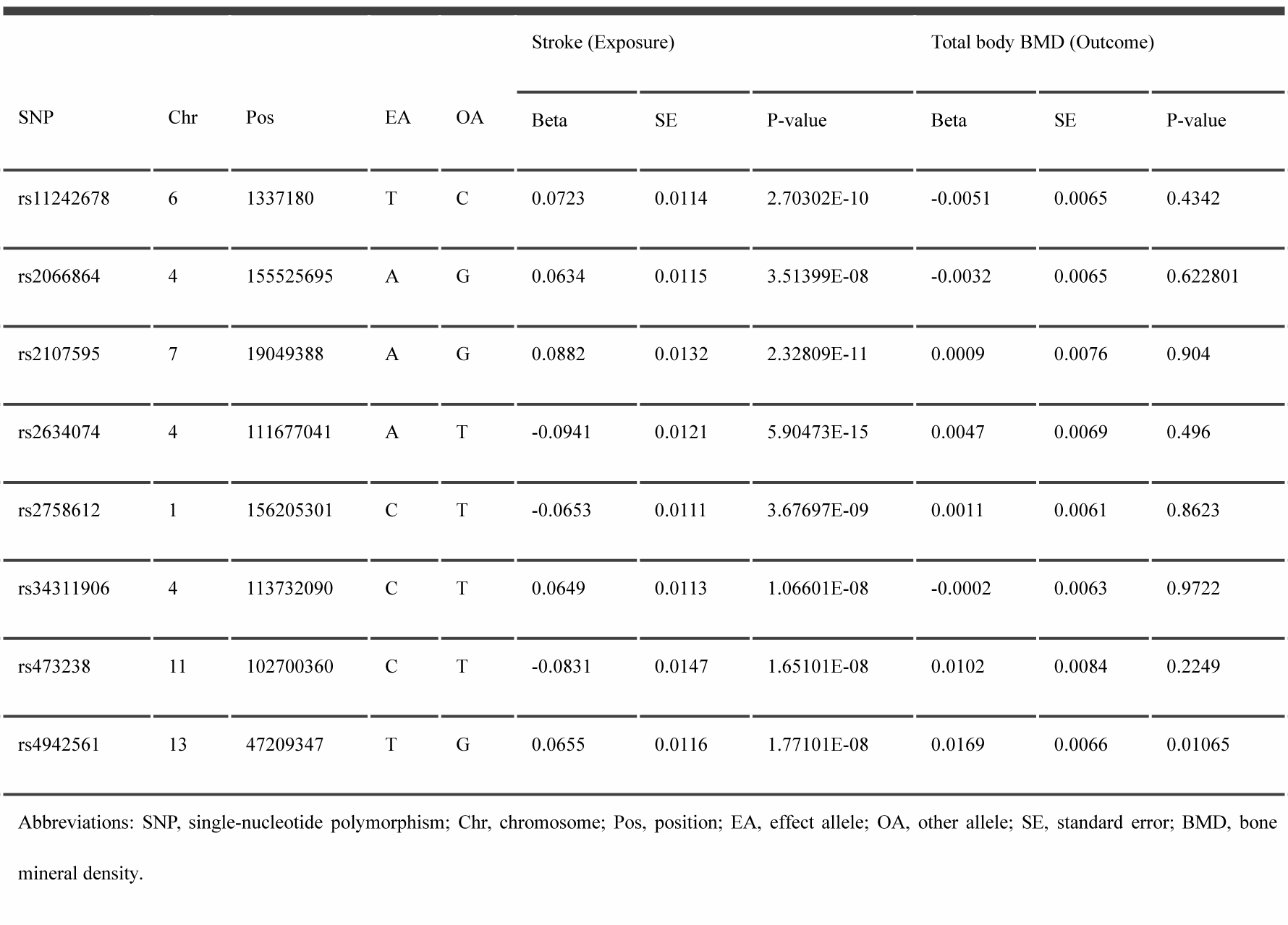
Association of the SNPs used as candidate genetic instruments from the GWAS for Mendelian randomization analyses of stroke and risk of total body BMD.

We evaluated the causal effect of CHD, HF and Stroke on total body BMD in the MR analysis (Table 7). CHD can significantly affect the incidence of total body BMD, and every higher standard deviation in the risk of CHD decreased the average total body BMD by 0.0459 units in the IVW analysis (Beta = -0.0459, 95%CI = -0.0815–-0.0104, P = 0.0113)(Table 7). The weighted median method and the MR-Egger method yielded non-significant results (Table 7). HF and Stroke showed no remarkable influence on total body BMD based on the IVW, the weighted median, the MR-Egger analyses (Table 7).

**Table 7.**
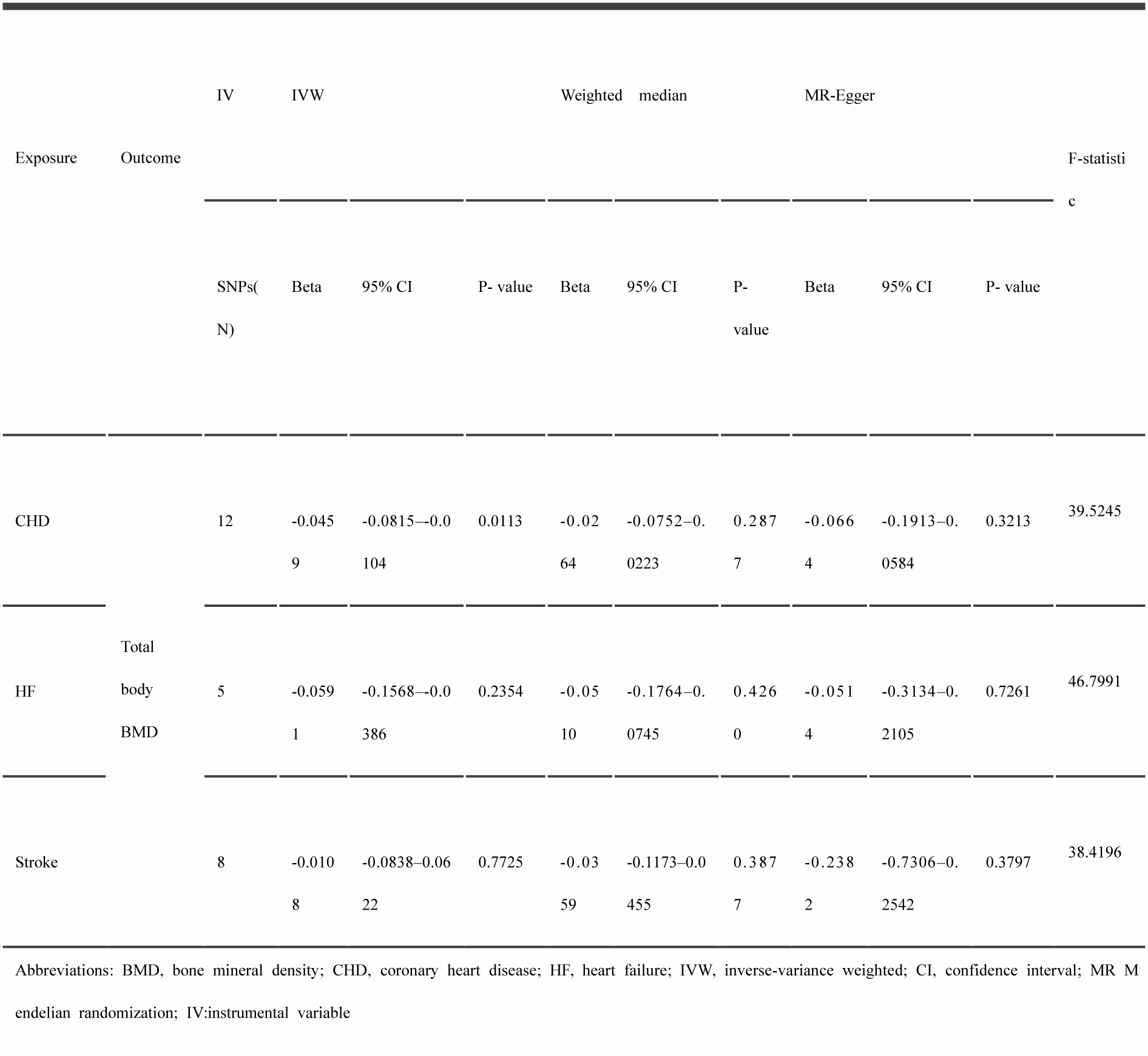
Mendelian Randomization estimates of CHD, HF and Stroke on total body BMD.

The estimated effect sizes for exposure (CHD, HF, stroke) and outcome total body BMD are shown in the scatter plot (Figure 2). The forest plot is shown in Figure 3.

**Figure 2.**
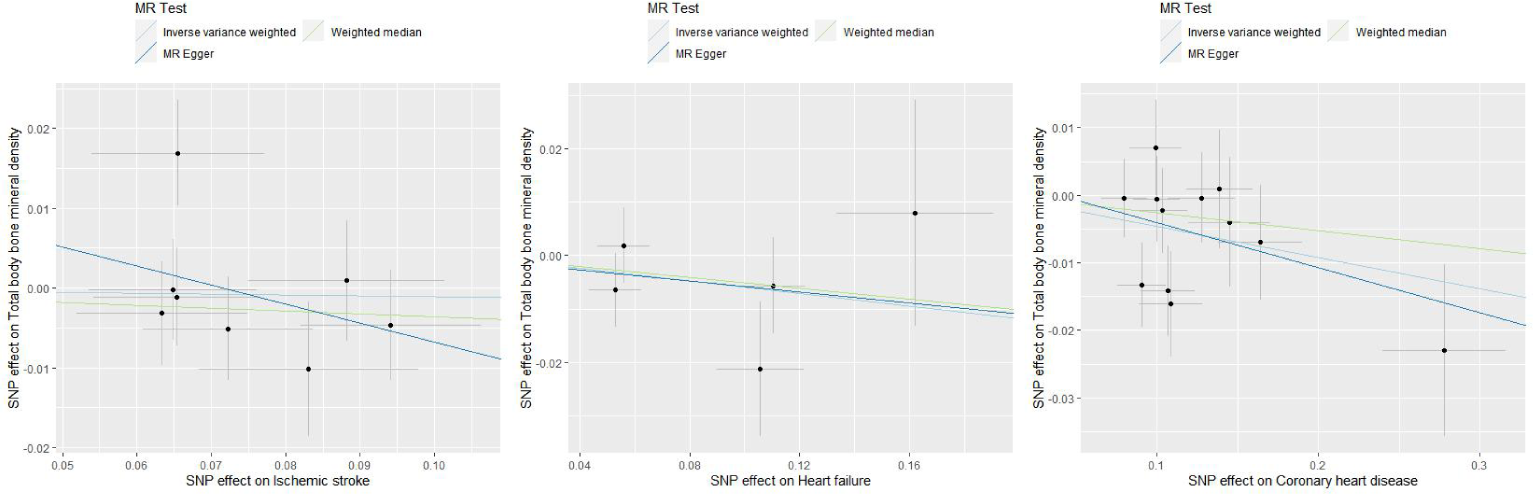
Scatter plots of estimated effect sizes for exposure (coronary heart disease, heart failure, stroke) on outcome (total body bone mineral density). ‘SNP’ denotes Single Nucleotide Polymorphism.

**Figure 3.**
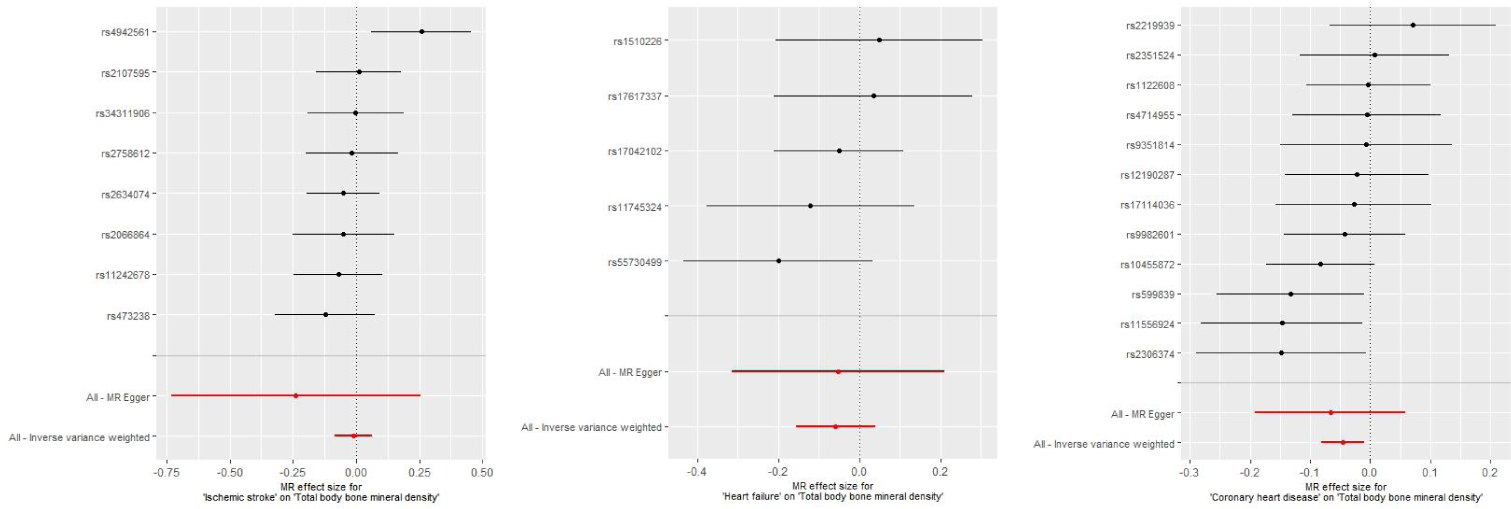
Forest plots of estimated effect sizes for exposure (coronary heart disease, heart failure, stroke) on outcome (total body bone mineral density). ‘MR’ refers to Mendelian randomization.

Heterogeneity tests revealed a lack of significant variation (P > 0.05) as indicated in Table 8. Additionally, the MR-Egger intercept suggested an absence of notable horizontal pleiotropy (intercept P > 0.05), detailed in Table 9. To evaluate the robustness of individual SNP effects, sensitivity was assessed using the leave-one-out strategy, depicted in Figure 4.

**Figure 4.**
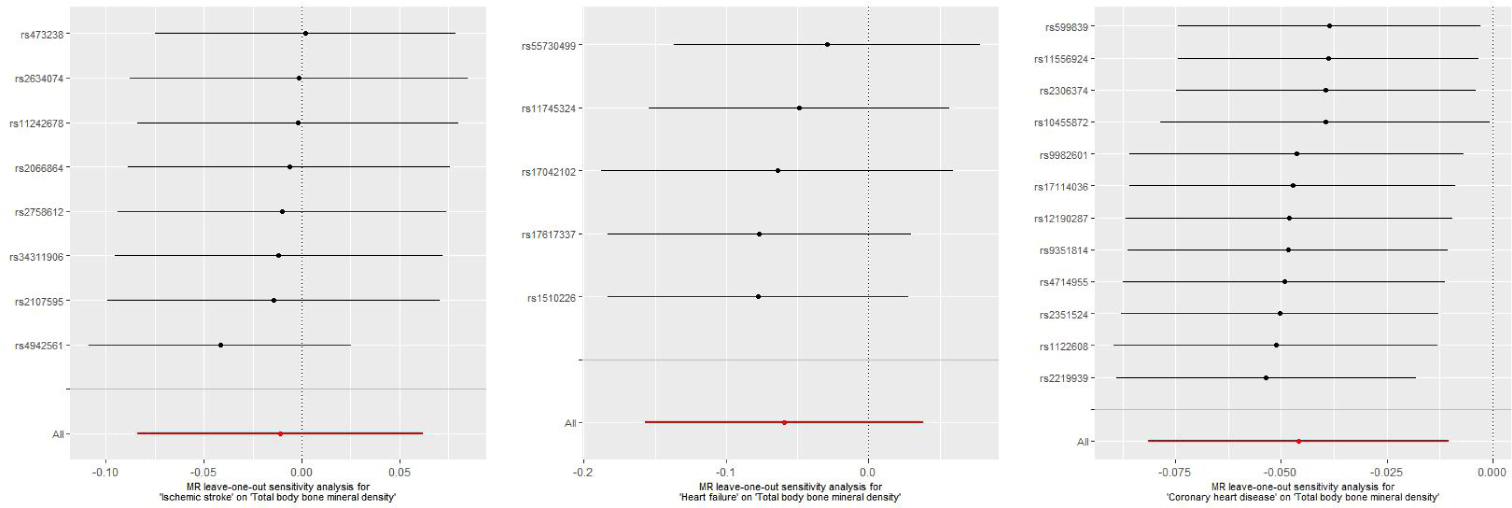
The leave-one-out method for exposure (coronary heart disease, heart failure, stroke) on outcome (total body bone mineral density). ‘MR’ refers to Mendelian randomization.

**Table 8.**
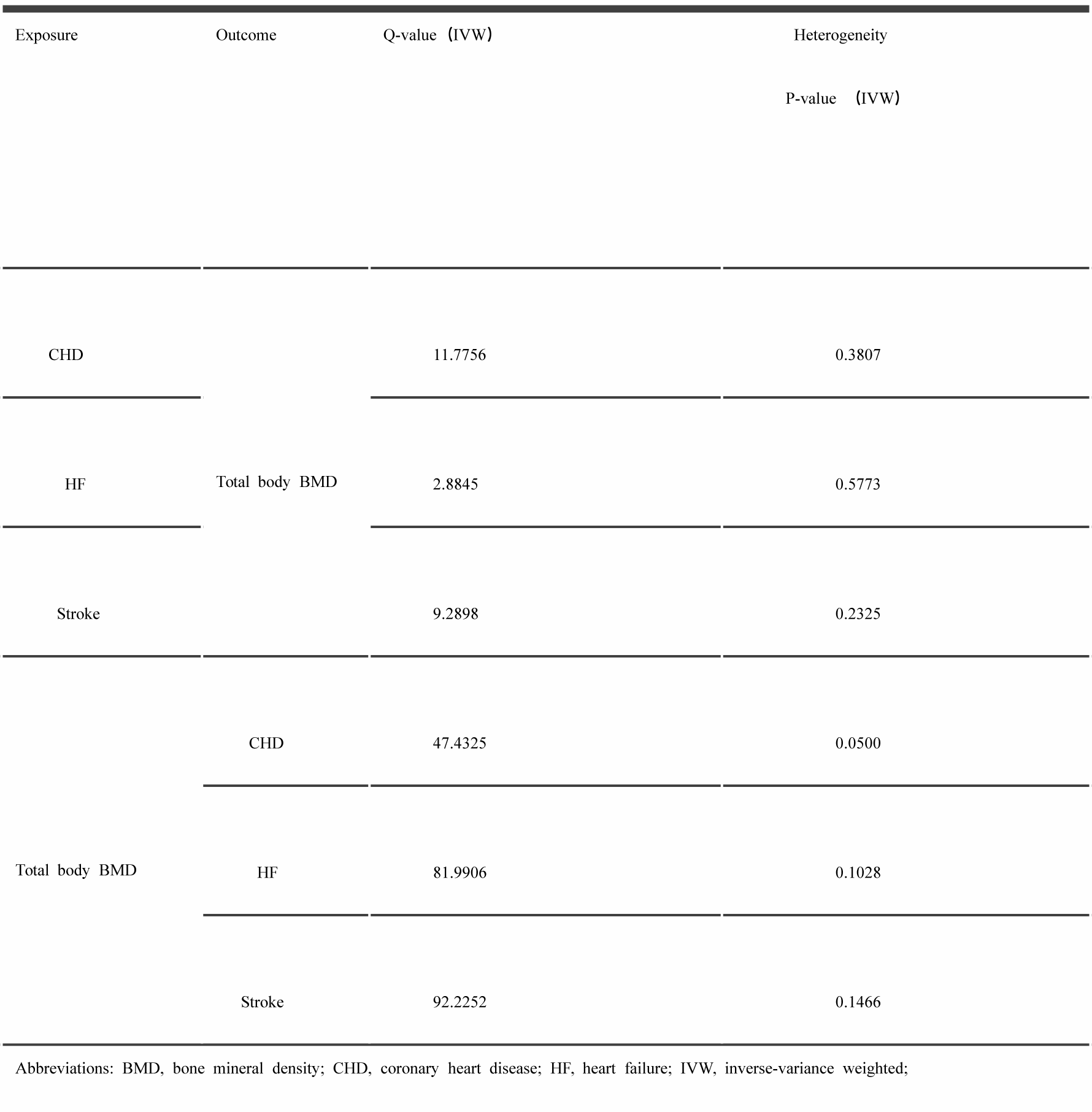
Results of heterogeneity analysis.

| Exposure | Outcome | Q-value (IVW) | Heterogeneity |
| --- | --- | --- | --- |
|  |  |  | P-value (IVW) |
| CHD |  | 11.7756 | 0.3807 |
| HF | Total body BMD | 2.8845 | 0.5773 |
| Stroke |  | 9.2898 | 0.2325 |
| Total body BMD | CHD | 47.4325 | 0.0500 |
|  | HF | 81.9906 | 0.1028 |
|  | Stroke | 92.2252 | 0.1466 |

Abbreviations: BMD, bone mineral density; CHD, coronary heart disease; HF, heart failure; IVW, inverse-variance weighted;

**Table 9.** Results of MR-Egger intercept test.

| Exposure | Outcome | Egger intercept | P-value | Se |
| --- | --- | --- | --- | --- |
| CHD | Total body BMD | 0.0025 | 0.7429 | 0.0076 |
| HF |  | -0.0007 | 0.9542 | 0.0107 |
| Stroke |  | 0.0170 | 0.3952 | 0.0186 |
| Total body BMD | CHD | -0.0057 | 0.4995 | 0.0084 |
|  | HF | 0.0014 | 0.6758 | 0.0033 |
|  | Stroke | -0.0029 | 0.4616 | 0.0039 |

Abbreviations: BMD, bone mineral density; CHD, coronary heart disease; HF, heart failure; Se, standard error

The presence of horizontal pleiotropy was further investigated using MR-PRESSO. This analysis identified a number of aberrant SNPs within the IVs concerning total body BMD on HF, as shown in Table 10. Upon the exclusion of these outliers, subsequent MR-PRESSO analyses did not detect any outlier influences, as reported in Table 11.

**Table 10.**
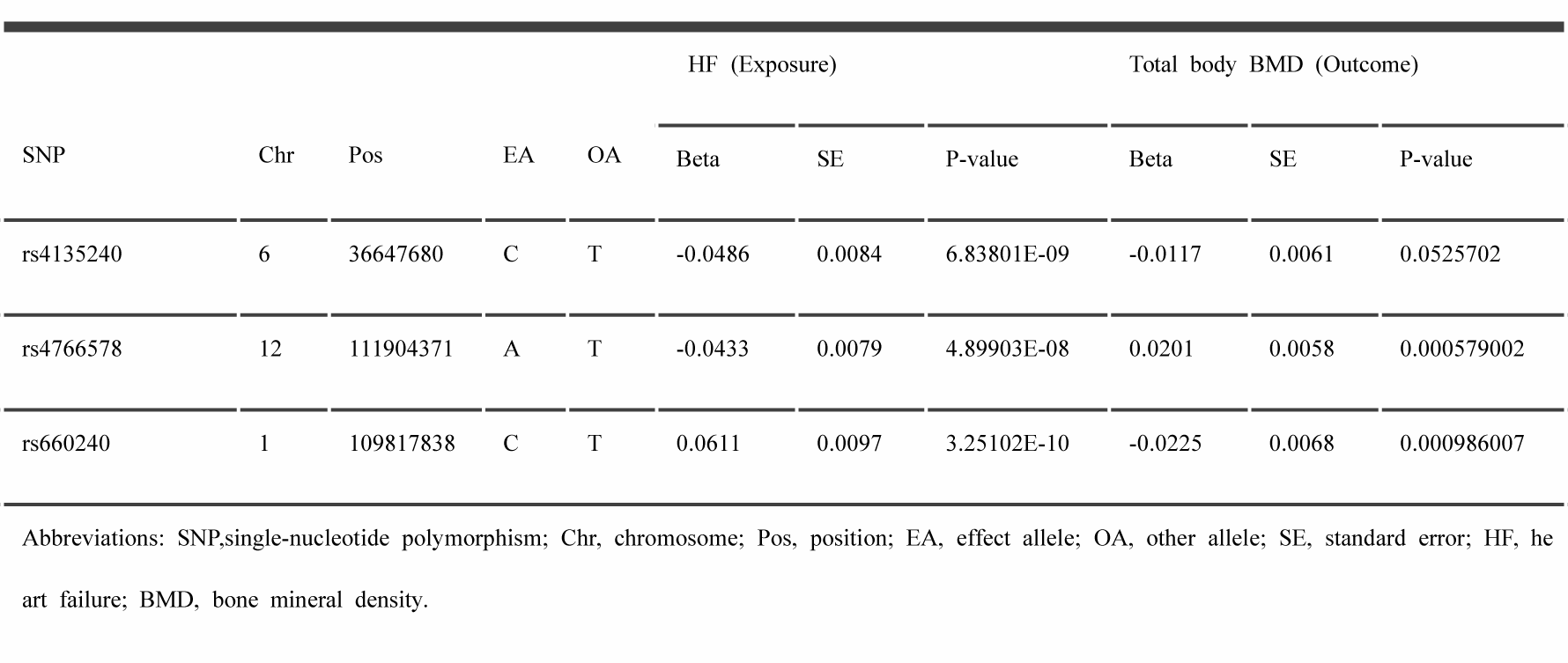
Abnormal SNPs in MR-PRESSO analysis of HF on total body BMD.

**Table 11.**
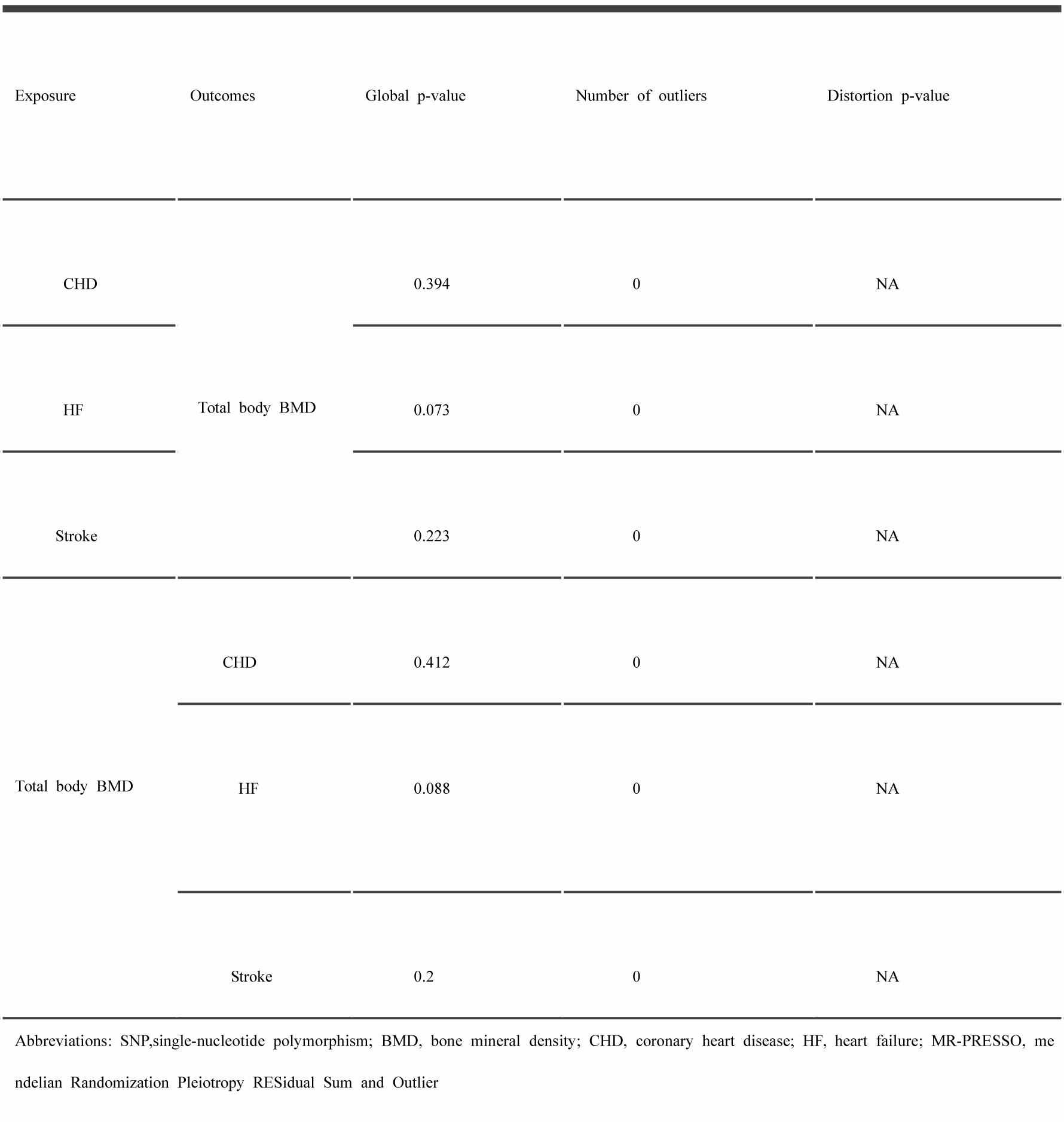
MR-PRESSO test of all the results.

### 2. Influence of total body BMD on CVD (CHD, HF, Stroke): A Causal Assessment

Confounding genes removed are shown in Table 12, 13, 14. Details used for the SNPs related to total body BMD and the SNPs associated with CVD (CHD, HF, stroke) are listed in Table 15, 16, 17.

**Table 12.** The removed SNP containing known confounding factors of total body BMD on CHD.

| SNP | Chr | Pos | EA | OA | Total body BMD (Exposure) |  |  | CHD (Outcome) |  |  |
| --- | --- | --- | --- | --- | --- | --- | --- | --- | --- | --- |
|  |  |  |  |  | Beta | SE | P-value | Beta | SE | P-value |
| rs13204965 | 6 | 127167072 | C | A | -0.0619 | 0.007 | 1.01508E-18 | 0.0137275 | 0.0170065 | 0.419557 |

Abbreviations: SNP, single-nucleotide polymorphism; Chr, chromosome; Pos, position; EA, effect allele; OA, other allele; SE, standard error; BMD, bone mineral density; CHD, coronary heart disease.

**Table 13.** The removed SNP containing known confounding factors of total body BMD on HF.

| SNP | Chr | Pos | EA | OA | Total body BMD (Exposure) |  |  | HF (Outcome) |  |  |
| --- | --- | --- | --- | --- | --- | --- | --- | --- | --- | --- |
|  |  |  |  |  | Beta | SE | P-value | Beta | SE | P-value |
| rs13204965 | 6 | 127167072 | C | A | -0.0619 | 0.007 | 1.01508E-18 | -0.0011 | 0.0092 | 0.9065 |
| rs780096 | 2 | 27741072 | G | C | 0.0311 | 0.0057 | 4.57699E-08 | -0.0079 | 0.0079 | 0.3157 |

Abbreviations: SNP, single-nucleotide polymorphism; Chr, chromosome; Pos, position; EA, effect allele; OA, other allele; SE, standard error; BMD, bone mineral density; HF, heart failure.

**Table 14.** The removed SNP containing known confounding factors of total body BMD on stroke.

| SNP | Chr | Pos | EA | OA | Total body BMD (Exposure) |  |  | Stroke (Outcome) |  |  |
| --- | --- | --- | --- | --- | --- | --- | --- | --- | --- | --- |
|  |  |  |  |  | Beta | SE | P-value | Beta | SE | P-value |
| rs13204965 | 6 | 127167072 | C | A | -0.0619 | 0.007 | 1.01508E-18 | -0.0286 | 0.012 | 0.01741 |
| rs780096 | 2 | 27741072 | G | C | 0.0311 | 0.0057 | 4.57699E-08 | -0.0113 | 0.0107 | 0.2926 |

Abbreviations: SNP, single-nucleotide polymorphism; Chr, chromosome; Pos, position; EA, effect allele; OA, other allele; SE, standard error; BMD, bone mineral density.

**Table 15.** Association of the SNPs used as candidate genetic instruments from the GWAS for Mendelian randomization analyses of total body BMD and risk of CHD.

| SNP | Chr | Pos | EA | OA | Total body BMD (Exposure) |  |  | CHD (Outcome) |  |  |
| --- | --- | --- | --- | --- | --- | --- | --- | --- | --- | --- |
|  |  |  |  |  | Beta | SE | P-value | Beta | SE | P-value |
| rs10490046 | 2 | 40630678 | C | A | -0.0429 | 0.0067 | 1.434E-10 | -0.0250067 | 0.0189856 | 0.187791 |
| rs10493013 | 1 | 22703035 | C | T | 0.1013 | 0.0074 | 4.07474E-43 | -0.0033062 | 0.0180114 | 0.854356 |
| rs10777212 | 12 | 90334829 | T | G | 0.0452 | 0.006 | 5.05126E-14 | 0.0155206 | 0.0149326 | 0.29863 |
| rs10788264 | 10 | 124015986 | A | G | -0.0338 | 0.0057 | 2.60597E-09 | 0.0298681 | 0.0144376 | 0.0385665 |
| rs10832520 | 11 | 15816918 | A | T | 0.1123 | 0.0158 | 1.00092E-12 | 0.0003941 | 0.0406626 | 0.992266 |
| rs11745493 | 5 | 122847622 | G | A | -0.0445 | 0.0065 | 7.74462E-12 | -0.0157347 | 0.0159937 | 0.32521 |
| rs11898505 | 2 | 54684557 | G | A | -0.0342 | 0.006 | 1.28201E-08 | 0.01481 | 0.0173628 | 0.393672 |
| rs11904127 | 2 | 85484818 | A | G | -0.0324 | 0.0057 | 1.18201E-08 | 0.0147723 | 0.0142359 | 0.299419 |
| rs11910328 | 21 | 40350744 | A | G | -0.0429 | 0.0077 | 2.99302E-08 | -0.0157575 | 0.0209145 | 0.451196 |
| rs11995824 | 8 | 120012700 | G | C | -0.0675 | 0.0058 | 1.06194E-31 | 0.009329 | 0.014028 | 0.506032 |
| rs1286150 | 14 | 91464890 | C | T | 0.0549 | 0.0072 | 2.44174E-14 | 0.0166644 | 0.0205679 | 0.417816 |
| rs1452102 | 21 | 28773868 | G | T | 0.0345 | 0.0057 | 1.736E-09 | 0.0147094 | 0.0141731 | 0.299344 |
| rs1548607 | 7 | 50901491 | G | A | -0.0363 | 0.0066 | 4.18398E-08 | -0.00224 | 0.0241312 | 0.926041 |
| rs2252865 | 1 | 8422676 | C | T | 0.0328 | 0.006 | 4.71998E-08 | -0.00592 | 0.0156861 | 0.705877 |
| rs2289410 | 2 | 42284110 | T | A | -0.0494 | 0.0088 | 2.00198E-08 | -0.01071 | 0.0203295 | 0.598319 |
| rs2350085 | 2 | 202799604 | C | T | 0.0643 | 0.0085 | 3.7949E-14 | -0.0028758 | 0.0258901 | 0.911555 |
| rs2553773 | 11 | 35083633 | G | C | 0.037 | 0.0058 | 1.493E-10 | -0.0050188 | 0.0140362 | 0.720671 |
| rs3743347 | 15 | 67547301 | A | C | 0.0519 | 0.0068 | 1.75186E-14 | 0.0026659 | 0.0166768 | 0.872992 |
| rs3801387 | 7 | 120974765 | G | A | 0.1347 | 0.0063 | 1.15001E-100 | 0.0058124 | 0.0164557 | 0.723929 |
| rs4757350 | 11 | 15703674 | T | C | -0.0564 | 0.0069 | 3.75405E-16 | -0.0332794 | 0.017658 | 0.0594758 |
| rs4846580 | 1 | 219897941 | A | G | 0.0345 | 0.0058 | 3.21299E-09 | 0.0011301 | 0.0140137 | 0.935728 |
| rs633995 | 1 | 172186729 | A | G | 0.0351 | 0.0058 | 1.61298E-09 | -0.0268112 | 0.0152785 | 0.0792885 |
| rs634277 | 11 | 86887931 | G | A | -0.0607 | 0.0061 | 2.15179E-23 | 0.0061239 | 0.0150167 | 0.683419 |
| rs6465511 | 7 | 96134115 | G | C | 0.0738 | 0.006 | 1.02802E-34 | 0.0428886 | 0.0151026 | 0.00451398 |
| rs6960249 | 7 | 96660132 | G | T | -0.0325 | 0.0057 | 1.44901E-08 | 0.0008693 | 0.0139862 | 0.95044 |
| rs725670 | 11 | 121913230 | A | G | -0.0322 | 0.0059 | 3.61202E-08 | -0.0105581 | 0.0150037 | 0.481621 |
| rs7586085 | 2 | 166577489 | G | A | -0.0532 | 0.0057 | 8.63575E-21 | 0.0080914 | 0.013737 | 0.555849 |
| rs7740042 | 6 | 151971720 | A | T | -0.0494 | 0.0071 | 2.70707E-12 | -0.0007803 | 0.0180184 | 0.965456 |
| rs7741085 | 6 | 44636919 | T | C | 0.0423 | 0.0057 | 1.50904E-13 | -0.0203963 | 0.0144119 | 0.156997 |
| rs8070128 | 17 | 17804725 | T | C | -0.0394 | 0.0059 | 1.98381E-11 | -0.0602312 | 0.0152666 | 7.97003E-05 |
| rs818427 | 5 | 112221869 | T | C | 0.0342 | 0.0061 | 2.37197E-08 | -0.0096668 | 0.0147448 | 0.512079 |
| rs884205 | 18 | 60054857 | C | A | 0.0531 | 0.0068 | 4.39036E-15 | -0.0069028 | 0.0209862 | 0.742215 |
| rs9594738 | 13 | 42952145 | T | C | -0.0614 | 0.0057 | 3.84061E-27 | -0.0026926 | 0.0138033 | 0.845337 |
| rs9910055 | 17 | 42283037 | T | C | 0.0442 | 0.0067 | 3.11889E-11 | -0.0244568 | 0.0168909 | 0.147638 |
| rs9972944 | 17 | 63771079 | G | A | -0.0363 | 0.0059 | 6.86594E-10 | -0.0024013 | 0.0143354 | 0.866968 |
| rs9976876 | 21 | 36970350 | T | G | -0.0375 | 0.0058 | 8.00571E-11 | 0.0051571 | 0.014061 | 0.713793 |

Abbreviations: SNP, single-nucleotide polymorphism; Chr, chromosome; Pos, position; EA, effect allele; OA, other allele; SE, standard error; BMD, bone mineral density; CHD, coronary heart disease.

**Table 16.** Association of the SNPs used as candidate genetic instruments from the GWAS for Mendelian randomization analyses of total body BM D and risk of HF.

| SNP | Chr | Pos | EA | OA | Total body BMD (Exposure) |  |  | HF (Outcome) |  |  |
| --- | --- | --- | --- | --- | --- | --- | --- | --- | --- | --- |
|  |  |  |  |  | Beta | SE | P-value | Beta | SE | P-value |
| rs10048745 | 2 | 68962137 | A | G | -0.0389 | 0.0067 | 6.44006E-09 | -0.0035 | 0.0092 | 0.7077 |
| rs1037011 | 12 | 107302778 | C | T | 0.0404 | 0.0057 | 1.54099E-12 | -0.0234 | 0.0078 | 0.00285397 |
| rs10490046 | 2 | 40630678 | C | A | -0.0429 | 0.0067 | 1.434E-10 | -0.0093 | 0.0094 | 0.3206 |
| rs10493013 | 1 | 22703035 | C | T | 0.1013 | 0.0074 | 4.07474E-43 | -0.002 | 0.0104 | 0.8496 |
| rs10735851 | 12 | 53743064 | A | G | -0.0541 | 0.0063 | 5.84252E-18 | -0.0175 | 0.0087 | 0.0439805 |
| rs10777212 | 12 | 90334829 | T | G | 0.0452 | 0.006 | 5.05126E-14 | 0.0025 | 0.0083 | 0.765799 |
| rs10788264 | 10 | 124015986 | A | G | -0.0338 | 0.0057 | 2.60597E-09 | 0.0122 | 0.0079 | 0.123 |
| rs10832520 | 11 | 15816918 | A | T | 0.1123 | 0.0158 | 1.00092E-12 | -0.0316 | 0.0231 | 0.1715 |
| rs10901216 | 9 | 133471891 | A | G | -0.0474 | 0.0061 | 5.5335E-15 | -0.0056 | 0.0082 | 0.4942 |
| rs10931982 | 2 | 202832130 | C | T | 0.0508 | 0.009 | 1.585E-08 | -0.0049 | 0.0108 | 0.649101 |
| rs11228240 | 11 | 68218290 | T | C | -0.083 | 0.0067 | 1.71791E-35 | -0.0026 | 0.0089 | 0.7724 |
| rs1159798 | 10 | 54412493 | C | A | -0.0429 | 0.007 | 1.014E-09 | 0.0142 | 0.0098 | 0.148 |
| rs11745493 | 5 | 122847622 | G | A | -0.0445 | 0.0065 | 7.74462E-12 | -0.0112 | 0.0091 | 0.2191 |
| rs117557198 | 12 | 49655948 | G | A | 0.0769 | 0.012 | 1.575E-10 | 0.0084 | 0.0153 | 0.585301 |
| rs11898505 | 2 | 54684557 | G | A | -0.0342 | 0.006 | 1.28201E-08 | 0.0038 | 0.0084 | 0.649299 |
| rs11904127 | 2 | 85484818 | A | G | -0.0324 | 0.0057 | 1.18201E-08 | -0.0008 | 0.0079 | 0.9226 |
| rs11910328 | 21 | 40350744 | A | G | -0.0429 | 0.0077 | 2.99302E-08 | 0.0133 | 0.0106 | 0.2119 |
| rs11934731 | 4 | 88831249 | A | G | -0.0674 | 0.0061 | 8.38687E-29 | 0.0038 | 0.0084 | 0.651199 |
| rs11995824 | 8 | 120012700 | G | C | -0.0675 | 0.0058 | 1.06194E-31 | -0.0033 | 0.0079 | 0.681 |
| rs12044944 | 1 | 240581653 | T | C | 0.0553 | 0.0074 | 7.54223E-14 | -0.0002 | 0.01 | 0.9816 |
| rs12258451 | 10 | 54423853 | G | C | -0.0702 | 0.0089 | 2.41213E-15 | -0.0041 | 0.0124 | 0.7431 |
| rs12442242 | 15 | 38340874 | G | A | 0.0509 | 0.0082 | 4.94106E-10 | 0.0234 | 0.0118 | 0.0469699 |
| rs12534510 | 7 | 120730944 | C | A | 0.0395 | 0.0057 | 3.14775E-12 | 0.01 | 0.0079 | 0.2059 |
| rs12612325 | 2 | 119632252 | A | G | -0.0548 | 0.0078 | 1.98198E-12 | -0.0062 | 0.0105 | 0.5562 |
| rs1286150 | 14 | 91464890 | C | T | 0.0549 | 0.0072 | 2.44174E-14 | -0.0125 | 0.0103 | 0.2271 |
| rs143187557 | 11 | 47284279 | T | C | -0.1237 | 0.0203 | 1.15401E-09 | -0.0106 | 0.028 | 0.704601 |
| rs144279715 | 2 | 119548256 | G | A | 0.2295 | 0.0294 | 6.18443E-15 | -0.0275 | 0.0323 | 0.3947 |
| rs144691710 | 17 | 41819562 | G | A | 0.1017 | 0.0113 | 2.23924E-19 | 0.0021 | 0.0145 | 0.8827 |
| rs1452102 | 21 | 28773868 | G | T | 0.0345 | 0.0057 | 1.736E-09 | -0.0039 | 0.0079 | 0.621199 |
| rs1548607 | 7 | 50901491 | G | A | -0.0363 | 0.0066 | 4.18398E-08 | 0.0044 | 0.0087 | 0.614301 |
| rs2252865 | 1 | 8422676 | C | T | 0.0328 | 0.006 | 4.71998E-08 | 0.0054 | 0.0083 | 0.5161 |
| rs2289410 | 2 | 42284110 | T | A | -0.0494 | 0.0088 | 2.00198E-08 | -0.0001 | 0.0121 | 0.9917 |
| rs2350085 | 2 | 202799604 | C | T | 0.0643 | 0.0085 | 3.7949E-14 | -0.0056 | 0.012 | 0.6389 |
| rs2414098 | 15 | 51537806 | C | T | 0.0329 | 0.0059 | 1.98701E-08 | -0.007 | 0.0119 | 0.554 |
| rs2553773 | 11 | 35083633 | G | C | 0.037 | 0.0058 | 1.493E-10 | 0.0119 | 0.0079 | 0.1288 |
| rs2566751 | 1 | 68664913 | A | T | -0.0567 | 0.01 | 1.31899E-08 | -0.024 | 0.0138 | 0.0815098 |
| rs2566752 | 1 | 68656697 | C | T | 0.0721 | 0.0059 | 1.87715E-34 | -0.0045 | 0.0081 | 0.5819 |
| rs34102936 | 7 | 38142840 | A | G | 0.0471 | 0.0057 | 1.86595E-16 | -0.0052 | 0.008 | 0.5181 |
| rs34670419 | 7 | 99130834 | T | G | -0.088 | 0.0154 | 1.087E-08 | -0.0551 | 0.0192 | 0.00421202 |
| rs35125553 | 12 | 1639249 | G | A | 0.0383 | 0.0066 | 5.202E-09 | -0.0038 | 0.0089 | 0.666099 |
| rs35199438 | 11 | 16630779 | T | G | -0.0489 | 0.0062 | 2.35993E-15 | 0.0035 | 0.0084 | 0.6758 |
| rs3743347 | 15 | 67547301 | A | C | 0.0519 | 0.0068 | 1.75186E-14 | -0.0208 | 0.0094 | 0.0260399 |
| rs3801387 | 7 | 120974765 | G | A | 0.1347 | 0.0063 | 1.15001E-100 | -0.0043 | 0.0089 | 0.6321 |
| rs447911 | 3 | 41127046 | C | G | 0.0708 | 0.0057 | 6.29361E-36 | 0.0002 | 0.0079 | 0.9813 |
| rs4757350 | 11 | 15703674 | T | C | -0.0564 | 0.0069 | 3.75405E-16 | 0.0106 | 0.0097 | 0.2714 |
| rs4846580 | 1 | 219897941 | A | G | 0.0345 | 0.0058 | 3.21299E-09 | 0.0065 | 0.0079 | 0.4136 |
| rs55781332 | 11 | 242859 | G | A | 0.0552 | 0.0069 | 8.07235E-16 | -0.0087 | 0.0093 | 0.349 |
| rs56104760 | 1 | 22486029 | G | A | -0.0747 | 0.0074 | 7.37734E-24 | 0.0131 | 0.0098 | 0.1801 |
| rs6029130 | 20 | 39103882 | T | C | 0.0348 | 0.0063 | 3.503E-08 | -0.0055 | 0.0089 | 0.5353 |
| rs6040063 | 20 | 10640877 | G | A | -0.0359 | 0.0056 | 1.78098E-10 | -0.0106 | 0.0078 | 0.175 |
| rs61884327 | 11 | 46766890 | C | T | 0.0801 | 0.0099 | 4.63447E-16 | 0.0143 | 0.0133 | 0.2806 |
| rs633995 | 1 | 172186729 | A | G | 0.0351 | 0.0058 | 1.61298E-09 | -0.0043 | 0.0081 | 0.5961 |
| rs634277 | 11 | 86887931 | G | A | -0.0607 | 0.0061 | 2.15179E-23 | 0.0052 | 0.0084 | 0.5389 |
| rs6465511 | 7 | 96134115 | G | C | 0.0738 | 0.006 | 1.02802E-34 | 0.0004 | 0.0082 | 0.9618 |
| rs6557155 | 6 | 151910126 | G | T | 0.0751 | 0.0059 | 2.55918E-37 | -0.0157 | 0.0081 | 0.0524699 |
| rs6960249 | 7 | 96660132 | G | T | -0.0325 | 0.0057 | 1.44901E-08 | -0.006 | 0.0079 | 0.4486 |
| rs7105860 | 11 | 27306364 | C | G | -0.0468 | 0.0059 | 2.35885E-15 | 0.0011 | 0.0081 | 0.8958 |
| rs71390846 | 16 | 86714715 | C | G | -0.0484 | 0.0075 | 1.38099E-10 | 0.0084 | 0.0101 | 0.4032 |
| rs725670 | 11 | 121913230 | A | G | -0.0322 | 0.0059 | 3.61202E-08 | -0.0027 | 0.0081 | 0.7392 |
| rs73169678 | 7 | 150953205 | A | C | 0.0619 | 0.0091 | 1.05293E-11 | -0.0191 | 0.0122 | 0.1172 |
| rs73305797 | 7 | 30997087 | T | A | -0.0422 | 0.0067 | 2.398E-10 | 0.01 | 0.0092 | 0.2759 |
| rs73719807 | 7 | 121191251 | C | A | 0.0925 | 0.0112 | 1.14288E-16 | -0.0252 | 0.0165 | 0.1263 |
| rs74394007 | 3 | 156692207 | C | A | -0.0608 | 0.0083 | 2.46093E-13 | -0.0004 | 0.0115 | 0.9742 |
| rs7548588 | 1 | 110475971 | C | T | 0.0367 | 0.0058 | 2.208E-10 | -0.0104 | 0.0079 | 0.1887 |
| rs757138 | 7 | 27989403 | G | T | 0.0348 | 0.0063 | 3.33403E-08 | 0.0048 | 0.0088 | 0.5875 |
| rs7586085 | 2 | 166577489 | G | A | -0.0532 | 0.0057 | 8.63575E-21 | -0.0004 | 0.0079 | 0.956 |
| rs76051363 | 4 | 1006987 | T | C | -0.0794 | 0.0085 | 1.38995E-20 | -0.0045 | 0.0113 | 0.691 |
| rs7728694 | 5 | 88288341 | T | G | -0.0503 | 0.0059 | 1.29599E-17 | -0.0125 | 0.008 | 0.1155 |
| rs7740042 | 6 | 151971720 | A | T | -0.0494 | 0.0071 | 2.70707E-12 | 0.0211 | 0.0098 | 0.03125 |
| rs7741085 | 6 | 44636919 | T | C | 0.0423 | 0.0057 | 1.50904E-13 | 0.0008 | 0.0079 | 0.9218 |
| rs78667121 | 13 | 43200103 | A | G | 0.1326 | 0.018 | 1.70098E-13 | 0.008 | 0.0231 | 0.728201 |
| rs8047501 | 16 | 392318 | G | A | -0.0524 | 0.0059 | 1.13397E-18 | 0.0044 | 0.0081 | 0.5894 |
| rs8070128 | 17 | 17804725 | T | C | -0.0394 | 0.0059 | 1.98381E-11 | -0.0223 | 0.008 | 0.00541203 |
| rs818427 | 5 | 112221869 | T | C | 0.0342 | 0.0061 | 2.37197E-08 | 0.0064 | 0.0082 | 0.4359 |
| rs838721 | 2 | 234303405 | G | A | 0.0314 | 0.0057 | 4.48002E-08 | -0.0118 | 0.0079 | 0.1326 |
| rs884205 | 18 | 60054857 | C | A | 0.0531 | 0.0068 | 4.39036E-15 | 0.0063 | 0.0091 | 0.489199 |
| rs9594738 | 13 | 42952145 | T | C | -0.0614 | 0.0057 | 3.84061E-27 | -0.0059 | 0.0079 | 0.4566 |
| rs9910055 | 17 | 42283037 | T | C | 0.0442 | 0.0067 | 3.11889E-11 | 0.004 | 0.0093 | 0.6693 |
| rs9972944 | 17 | 63771079 | G | A | -0.0363 | 0.0059 | 6.86594E-10 | 0.0018 | 0.0118 | 0.881 |
| rs9976876 | 21 | 36970350 | T | G | -0.0375 | 0.0058 | 8.00571E-11 | -0.0042 | 0.0079 | 0.5894 |

Abbreviations: SNP, single-nucleotide polymorphism; Chr, chromosome; Pos, position; EA, effect allele; OA, other allele; SE, standard error; BMD, bone mineral density; HF, heart failure.

**Table 17.** Association of the SNPs used as candidate genetic instruments from the GWAS for Mendelian randomization analyses of total body BM D and risk of stroke.

| SNP | Chr | Pos | EA | OA | Total body BMD (Exposure) |  |  | Stroke (Outcome) |  |  |
| --- | --- | --- | --- | --- | --- | --- | --- | --- | --- | --- |
|  |  |  |  |  | Beta | SE | P-value | Beta | SE | P-value |
| rs10048745 | 2 | 68962137 | A | G | -0.0389 | 0.0067 | 6.44006E-09 | -0.0087 | 0.0129 | 0.501601 |
| rs1037011 | 12 | 107302778 | C | T | 0.0404 | 0.0057 | 1.54099E-12 | 0.0019 | 0.0099 | 0.8492 |
| rs10490046 | 2 | 40630678 | C | A | -0.0429 | 0.0067 | 1.434E-10 | 0.0022 | 0.0126 | 0.8586 |
| rs10493013 | 1 | 22703035 | C | T | 0.1013 | 0.0074 | 4.07474E-43 | -0.0177 | 0.0131 | 0.1759 |
| rs10735851 | 12 | 53743064 | A | G | -0.0541 | 0.0063 | 5.84252E-18 | -0.0017 | 0.0103 | 0.8706 |
| rs10777212 | 12 | 90334829 | T | G | 0.0452 | 0.006 | 5.05126E-14 | -0.0066 | 0.0105 | 0.5308 |
| rs10788264 | 10 | 124015986 | A | G | -0.0338 | 0.0057 | 2.60597E-09 | 0.009 | 0.0099 | 0.3609 |
| rs10832520 | 11 | 15816918 | A | T | 0.1123 | 0.0158 | 1.00092E-12 | 0.031 | 0.0294 | 0.2922 |
| rs10901216 | 9 | 133471891 | A | G | -0.0474 | 0.0061 | 5.5335E-15 | -0.0013 | 0.0105 | 0.8999 |
| rs10931982 | 2 | 202832130 | C | T | 0.0508 | 0.009 | 1.585E-08 | 0.026 | 0.0153 | 0.0886605 |
| rs11228240 | 11 | 68218290 | T | C | -0.083 | 0.0067 | 1.71791E-35 | -0.0205 | 0.0118 | 0.0834392 |
| rs1159798 | 10 | 54412493 | C | A | -0.0429 | 0.007 | 1.014E-09 | -0.0008 | 0.0125 | 0.9519 |
| rs11745493 | 5 | 122847622 | G | A | -0.0445 | 0.0065 | 7.74462E-12 | 0.0159 | 0.0112 | 0.1586 |
| rs117557198 | 12 | 49655948 | G | A | 0.0769 | 0.012 | 1.575E-10 | 0.009 | 0.02 | 0.6543 |
| rs118115924 | 12 | 49379537 | T | G | -0.2822 | 0.0301 | 6.98554E-21 | 0.0487 | 0.0598 | 0.4154 |
| rs11898505 | 2 | 54684557 | G | A | -0.0342 | 0.006 | 1.28201E-08 | 0.0313 | 0.0107 | 0.00360296 |
| rs11904127 | 2 | 85484818 | A | G | -0.0324 | 0.0057 | 1.18201E-08 | -0.0197 | 0.0101 | 0.0498196 |
| rs11910328 | 21 | 40350744 | A | G | -0.0429 | 0.0077 | 2.99302E-08 | -0.0014 | 0.0135 | 0.9151 |
| rs11934731 | 4 | 88831249 | A | G | -0.0674 | 0.0061 | 8.38687E-29 | 0.0181 | 0.0106 | 0.0888198 |
| rs11995824 | 8 | 120012700 | G | C | -0.0675 | 0.0058 | 1.06194E-31 | 0.0021 | 0.0102 | 0.8398 |
| rs12044944 | 1 | 240581653 | T | C | 0.0553 | 0.0074 | 7.54223E-14 | -0.0061 | 0.0129 | 0.637699 |
| rs12258451 | 10 | 54423853 | G | C | -0.0702 | 0.0089 | 2.41213E-15 | -0.0058 | 0.0163 | 0.723199 |
| rs12442242 | 15 | 38340874 | G | A | 0.0509 | 0.0082 | 4.94106E-10 | 0.0082 | 0.0147 | 0.5779 |
| rs12534510 | 7 | 120730944 | C | A | 0.0395 | 0.0057 | 3.14775E-12 | 0.0058 | 0.0099 | 0.5595 |
| rs12612325 | 2 | 119632252 | A | G | -0.0548 | 0.0078 | 1.98198E-12 | -0.005 | 0.0142 | 0.7248 |
| rs1286150 | 14 | 91464890 | C | T | 0.0549 | 0.0072 | 2.44174E-14 | -0.0079 | 0.0131 | 0.5474 |
| rs143187557 | 11 | 47284279 | T | C | -0.1237 | 0.0203 | 1.15401E-09 | -0.0561 | 0.0374 | 0.1336 |
| rs144279715 | 2 | 119548256 | G | A | 0.2295 | 0.0294 | 6.18443E-15 | 0.0161 | 0.0508 | 0.7516 |
| rs144691710 | 17 | 41819562 | G | A | 0.1017 | 0.0113 | 2.23924E-19 | 0.0065 | 0.0197 | 0.7431 |
| rs1452102 | 21 | 28773868 | G | T | 0.0345 | 0.0057 | 1.736E-09 | -0.0033 | 0.0103 | 0.7461 |
| rs1548607 | 7 | 50901491 | G | A | -0.0363 | 0.0066 | 4.18398E-08 | 0.0012 | 0.0114 | 0.9181 |
| rs2252865 | 1 | 8422676 | C | T | 0.0328 | 0.006 | 4.71998E-08 | -0.0171 | 0.0104 | 0.099211 |
| rs2289410 | 2 | 42284110 | T | A | -0.0494 | 0.0088 | 2.00198E-08 | 0.0267 | 0.0162 | 0.0994008 |
| rs2350085 | 2 | 202799604 | C | T | 0.0643 | 0.0085 | 3.7949E-14 | 0.0144 | 0.0152 | 0.3436 |
| rs2414098 | 15 | 51537806 | C | T | 0.0329 | 0.0059 | 1.98701E-08 | 0.0205 | 0.0106 | 0.0538096 |
| rs2553773 | 11 | 35083633 | G | C | 0.037 | 0.0058 | 1.493E-10 | 0.0034 | 0.01 | 0.734999 |
| rs2566751 | 1 | 68664913 | A | T | -0.0567 | 0.01 | 1.31899E-08 | 0.0178 | 0.0182 | 0.3278 |
| rs2566752 | 1 | 68656697 | C | T | 0.0721 | 0.0059 | 1.87715E-34 | -0.0081 | 0.0102 | 0.4277 |
| rs2873195 | 17 | 2064702 | T | A | 0.0406 | 0.0062 | 4.31122E-11 | 0.021 | 0.0112 | 0.0601395 |
| rs34102936 | 7 | 38142840 | A | G | 0.0471 | 0.0057 | 1.86595E-16 | 0.0042 | 0.0098 | 0.668199 |
| rs34670419 | 7 | 99130834 | T | G | -0.088 | 0.0154 | 1.087E-08 | -0.0362 | 0.0258 | 0.1606 |
| rs35125553 | 12 | 1639249 | G | A | 0.0383 | 0.0066 | 5.202E-09 | 0.0157 | 0.0115 | 0.172 |
| rs35199438 | 11 | 16630779 | T | G | -0.0489 | 0.0062 | 2.35993E-15 | 0.0061 | 0.0106 | 0.569 |
| rs3743347 | 15 | 67547301 | A | C | 0.0519 | 0.0068 | 1.75186E-14 | -0.0103 | 0.012 | 0.3926 |
| rs3801387 | 7 | 120974765 | G | A | 0.1347 | 0.0063 | 1.15001E-100 | 0.0085 | 0.0111 | 0.4434 |
| rs447911 | 3 | 41127046 | C | G | 0.0708 | 0.0057 | 6.29361E-36 | 0.0039 | 0.0101 | 0.7014 |
| rs4757350 | 11 | 15703674 | T | C | -0.0564 | 0.0069 | 3.75405E-16 | -0.016 | 0.014 | 0.2511 |
| rs4846580 | 1 | 219897941 | A | G | 0.0345 | 0.0058 | 3.21299E-09 | -0.0077 | 0.0101 | 0.4448 |
| rs55781332 | 11 | 242859 | G | A | 0.0552 | 0.0069 | 8.07235E-16 | -0.0022 | 0.0119 | 0.8551 |
| rs56104760 | 1 | 22486029 | G | A | -0.0747 | 0.0074 | 7.37734E-24 | -0.0048 | 0.0128 | 0.709099 |
| rs6029130 | 20 | 39103882 | T | C | 0.0348 | 0.0063 | 3.503E-08 | 0.0181 | 0.0115 | 0.115 |
| rs6040063 | 20 | 10640877 | G | A | -0.0359 | 0.0056 | 1.78098E-10 | 0.002 | 0.0099 | 0.8378 |
| rs61884327 | 11 | 46766890 | C | T | 0.0801 | 0.0099 | 4.63447E-16 | 0.0334 | 0.0171 | 0.0509096 |
| rs633995 | 1 | 172186729 | A | G | 0.0351 | 0.0058 | 1.61298E-09 | -0.0105 | 0.01 | 0.294 |
| rs634277 | 11 | 86887931 | G | A | -0.0607 | 0.0061 | 2.15179E-23 | -0.011 | 0.0107 | 0.3026 |
| rs6465511 | 7 | 96134115 | G | C | 0.0738 | 0.006 | 1.02802E-34 | 0.016 | 0.0107 | 0.1363 |
| rs6557155 | 6 | 151910126 | G | T | 0.0751 | 0.0059 | 2.55918E-37 | -0.0141 | 0.0104 | 0.1756 |
| rs6960249 | 7 | 96660132 | G | T | -0.0325 | 0.0057 | 1.44901E-08 | 0.0076 | 0.0101 | 0.4499 |
| rs7105860 | 11 | 27306364 | C | G | -0.0468 | 0.0059 | 2.35885E-15 | 0.0165 | 0.0102 | 0.1056 |
| rs71390846 | 16 | 86714715 | C | G | -0.0484 | 0.0075 | 1.38099E-10 | -0.0235 | 0.013 | 0.0696194 |
| rs725670 | 11 | 121913230 | A | G | -0.0322 | 0.0059 | 3.61202E-08 | -0.0197 | 0.0106 | 0.0629898 |
| rs73169678 | 7 | 150953205 | A | C | 0.0619 | 0.0091 | 1.05293E-11 | -0.0014 | 0.0158 | 0.9283 |
| rs73305797 | 7 | 30997087 | T | A | -0.0422 | 0.0067 | 2.398E-10 | 0.0181 | 0.0118 | 0.1233 |
| rs73349318 | 10 | 112245400 | T | A | 0.0472 | 0.0085 | 2.68201E-08 | 0.0006 | 0.0157 | 0.9702 |
| rs73719807 | 7 | 121191251 | C | A | 0.0925 | 0.0112 | 1.14288E-16 | 0.007 | 0.0205 | 0.7329 |
| rs74394007 | 3 | 156692207 | C | A | -0.0608 | 0.0083 | 2.46093E-13 | 0.0061 | 0.0146 | 0.676 |
| rs7548588 | 1 | 110475971 | C | T | 0.0367 | 0.0058 | 2.208E-10 | 0.0003 | 0.0101 | 0.9753 |
| rs757138 | 7 | 27989403 | G | T | 0.0348 | 0.0063 | 3.33403E-08 | -0.0041 | 0.0114 | 0.721601 |
| rs7586085 | 2 | 166577489 | G | A | -0.0532 | 0.0057 | 8.63575E-21 | -0.0081 | 0.0099 | 0.4094 |
| rs76051363 | 4 | 1006987 | T | C | -0.0794 | 0.0085 | 1.38995E-20 | -0.013 | 0.015 | 0.3863 |
| rs7728694 | 5 | 88288341 | T | G | -0.0503 | 0.0059 | 1.29599E-17 | -0.0082 | 0.0103 | 0.4262 |
| rs7740042 | 6 | 151971720 | A | T | -0.0494 | 0.0071 | 2.70707E-12 | -0.0205 | 0.0126 | 0.1026 |
| rs7741085 | 6 | 44636919 | T | C | 0.0423 | 0.0057 | 1.50904E-13 | 0.003 | 0.0103 | 0.770699 |
| rs78667121 | 13 | 43200103 | A | G | 0.1326 | 0.018 | 1.70098E-13 | 0.0027 | 0.0315 | 0.9319 |
| rs8047501 | 16 | 392318 | G | A | -0.0524 | 0.0059 | 1.13397E-18 | 0.0086 | 0.0104 | 0.4088 |
| rs8070128 | 17 | 17804725 | T | C | -0.0394 | 0.0059 | 1.98381E-11 | -0.0163 | 0.0102 | 0.1088 |
| rs818427 | 5 | 112221869 | T | C | 0.0342 | 0.0061 | 2.37197E-08 | -0.0041 | 0.0105 | 0.6936 |
| rs838721 | 2 | 234303405 | G | A | 0.0314 | 0.0057 | 4.48002E-08 | -0.0025 | 0.0101 | 0.8065 |
| rs884205 | 18 | 60054857 | C | A | 0.0531 | 0.0068 | 4.39036E-15 | -0.0116 | 0.0119 | 0.3296 |
| rs9594738 | 13 | 42952145 | T | C | -0.0614 | 0.0057 | 3.84061E-27 | -0.0017 | 0.01 | 0.8612 |
| rs9910055 | 17 | 42283037 | T | C | 0.0442 | 0.0067 | 3.11889E-11 | 0.0032 | 0.0117 | 0.785899 |
| rs9972944 | 17 | 63771079 | G | A | -0.0363 | 0.0059 | 6.86594E-10 | -0.005 | 0.0103 | 0.6269 |
| rs9976876 | 21 | 36970350 | T | G | -0.0375 | 0.0058 | 8.00571E-11 | 0.0161 | 0.01 | 0.106 |

Abbreviations: SNP, single-nucleotide polymorphism; Chr, chromosome; Pos, position; EA, effect allele; OA, other allele; SE, standard error; BMD, bone mineral density.

We evaluated the causal effect of total body BMD on CVD (CHD, HF, stroke) in the MR analysis (Table 18). Total body BMD showed no remarkable influence on CHD based on the IVW analyses (Beta = 0.0651; 95%CI = -0.0606–0.1907, P = 0.3101). Total body BMD showed no remarkable influence on HF based on the IVW analyses (Beta =-0.0154; 95%CI = -0.0603–0.0295, P = 0.5014). Total body BMD showed no remarkable influence on Stroke based on the IVW analyses(Beta = 0.0237; 95% CI -0.0287–0.0762, P = 0.3754). The weighted median method and the MR-Egger method also indicated non-significant findings (Table 18).

**Table 18.**
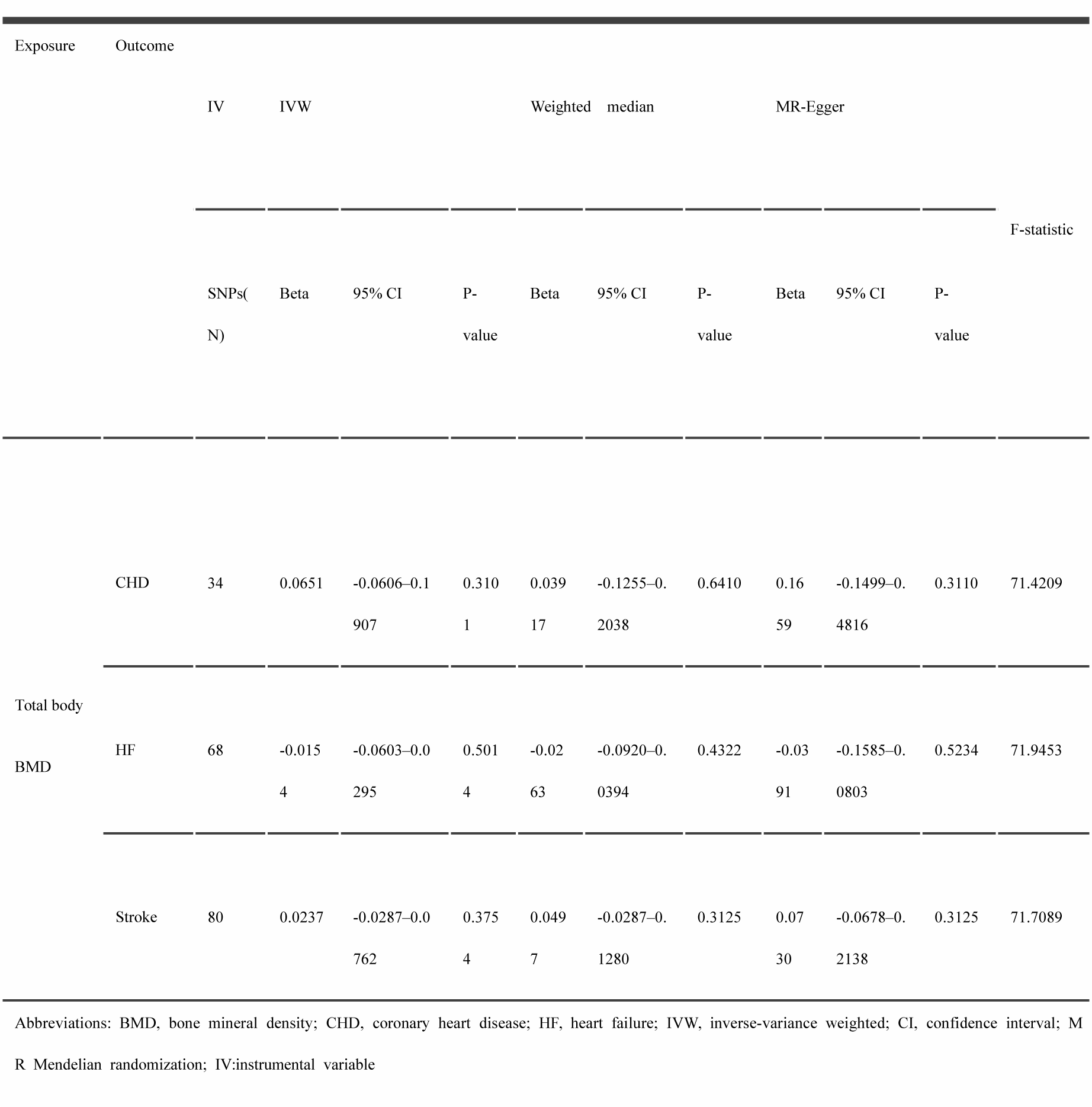
Mendelian Randomization estimates of CHD, HF and stroke on total body BMD.

Scatter plot representations of the estimated effect sizes between total body BMD and CVD (CHD, HF, Stroke) are depicted in Figure 5, with a corresponding forest plot presented in Figure 6.

**Figure 5.**
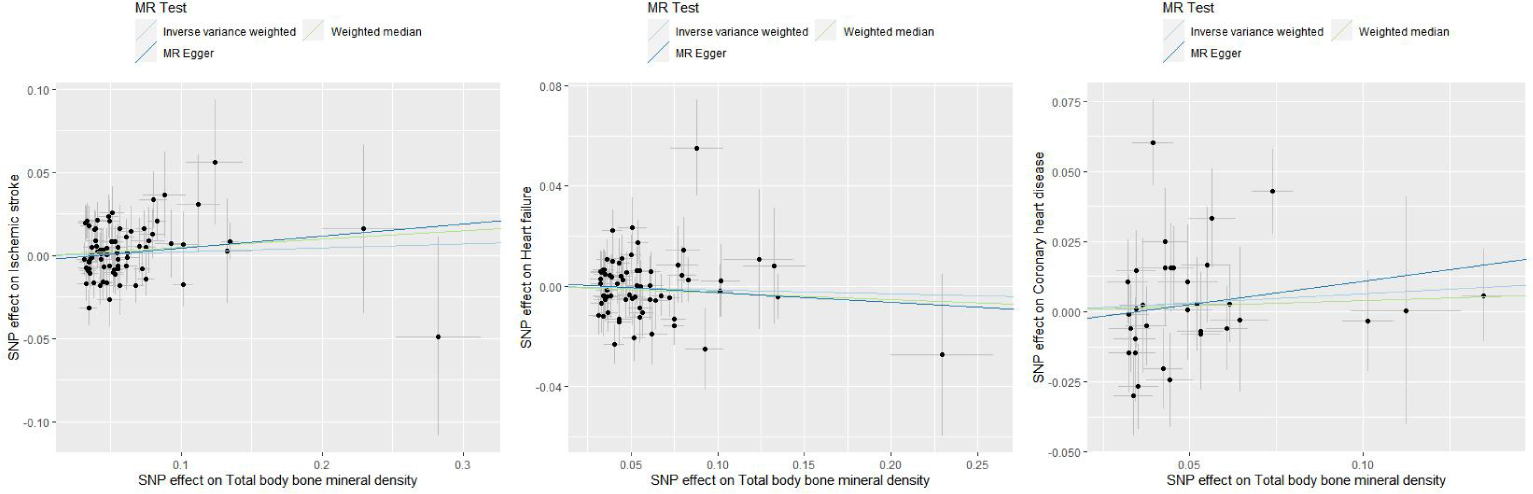
Scatter plots of estimated effect sizes for exposure (total body bone mineral density) on outcome (coronary heart disease, heart failure, stroke). ‘SNP’ denotes Single Nucleotide Polymorphism.

**Figure 6.**
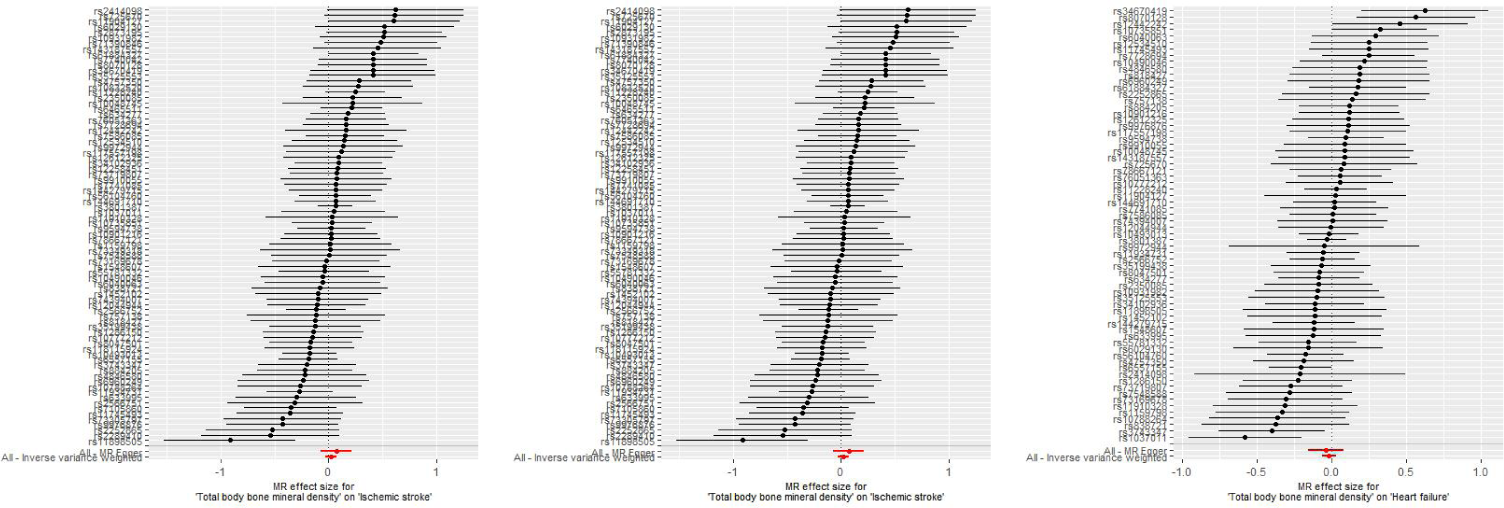
Forest plots of estimated effect sizes for exposure (total body bone mineral density) on outcome (coronary heart disease, heart failure, stroke). ‘MR’ refers to Mendelian randomization.

Heterogeneity assessments indicated no significant discrepancies (P > 0.05), as recorded in Table 8. Likewise, evidence for horizontal pleiotropy was not observed in the MR-Egger intercept results (intercept P > 0.05), listed in Table 9.

The influence of individual SNPs was scrutinized using the leave-one-out approach as a part of our sensitivity analysis, the results of which are displayed in Figure 7.

**Figure 7.**
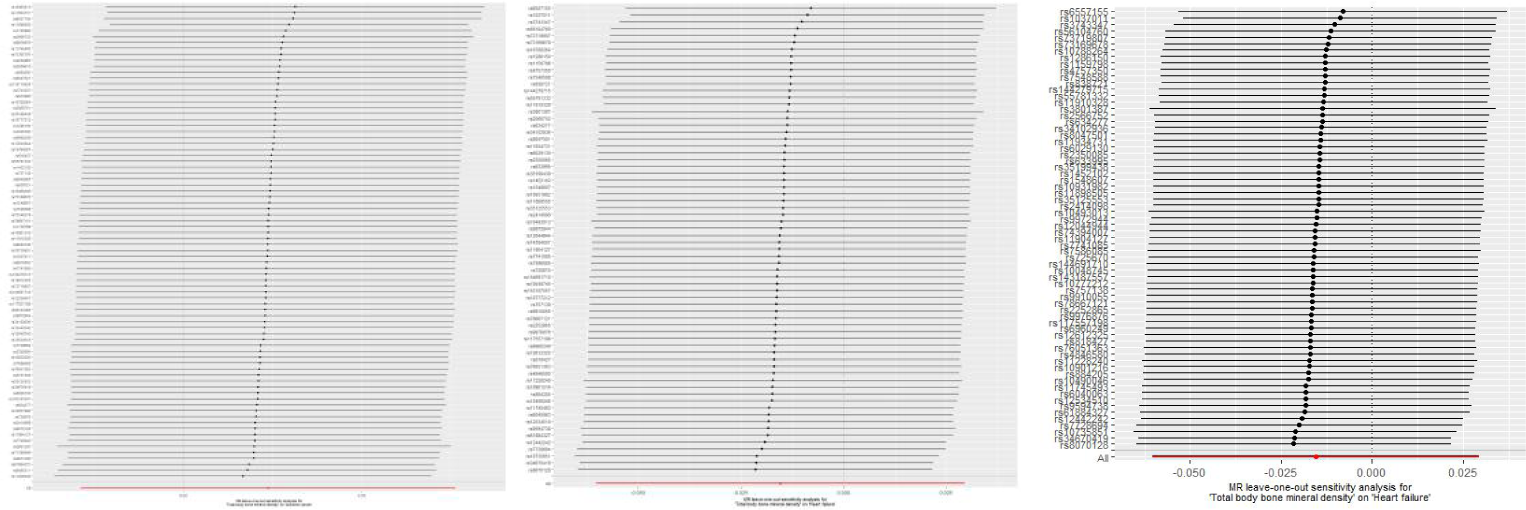
The leave-one-out method for exposure (total body bone mineral density) on outcome (coronary heart disease, heart failure, stroke). ‘MR’ refers to Mendelian randomization.

Horizontal pleiotropy was further examined via MR-PRESSO analysis. This analysis identified several atypical SNPs within the IVs for total body BMD’s influence on HF, detailed in Table 19. Subsequent to outlier removal, no further MR-PRESSO outlier detections were reported, as evidenced in Table 11.

**Table 19.** Abnormal SNPs in MR-PRESSO analysis of total body BMD on HF.

| SNP | Chr | Pos | EA | OA | Total body BMD (Exposure) |  |  | HF (Outcome) |  |  |
| --- | --- | --- | --- | --- | --- | --- | --- | --- | --- | --- |
|  |  |  |  |  | Beta | SE | P-value | Beta | SE | P-value |
| rs2873195 | 17 | 2064702 | T | A | 0.0406 | 0.0062 | 4.31122E-11 | 0.0377 | 0.0123 | 0.00226798 |

Abbreviations: SNP, single-nucleotide polymorphism; Chr, chromosome; Pos, position; EA, effect allele; OA, other allele; SE, standard error; HF, heart failure; BMD, bone mineral density.
#

## V. Discussion

This study represents the most extensive MR investigation into the link between CVD and the risk of OP. The findings from our MR analysis suggest that individuals with CHD may have an increased risk of developing OP. Additionally, the association persisted following sensitivity testing, with no single instrumental variable substantially altering the result.

While the exact pathways driving the relationship between CHD and OP risk remain to be elucidated, there is an accumulating body of research pointing towards a correlation between the two. Numerous large-scale epidemiological studies have identified an association between CHD and OP.

### 1. Shared Etiological Influences

The hypothesis posits that the concurrent presence of CHD and OP may arise from shared causal factors, such as hypertension, alcohol consumption, physical inactivity, and smoking, which may simultaneously contribute to both atherosclerosis and bone loss. This might shed light on the partial linkage observed between these two conditions^33^. Nevertheless, the link between CHD and OP has been consistently observed in numerous epidemiological studies, even after adjusting for many of these risk factors.

### 2. Common pathophysiological mechanisms

Approximately 90% of coronary atherosclerosis cases are accompanied by coronary artery calcification. It was previously believed that vascular calcification was simply the deposition of calcium in local vascular areas. However, recent research has indicated that vascular calcification is not merely a passive accumulation of calcium salts. Instead, it involves the transformation of local vascular cells into osteogenic cells, and local vascular tissue assumes characteristics akin to bone tissue. This is a highly regulated and reversible biological process, which bears significant similarities to the process of bone mineralization^34^. Vascular wall cells, including fibroblasts, endothelial cells, macrophages, and vascular smooth muscle cells — particularly the latter under the influence of inflammatory factors and transforming growth factor beta (TGF-β) — can differentiate into osteogenic cells. These osteogenic cells can then secrete a variety of proteins related to bone formation, such as bone morphogenic protein and osteopontin, which may underlie the association between localized vascular calcification and bone metabolism. Based on this association, some scholars have proposed the concept of the ‘bone-vascular axis’ and posit that vascular calcification is, in essence, an intravascular, programmed osteogenic process driven by various factors^35^. The similarities and connections between the bone metabolic process and the vascular calcification process may explain the link between CHD and OP. The current study suggests that the common molecular and biological mechanisms underlying the pathogenesis of CHD and OP, along with the biochemical factors impacting both, may primarily include aspects such as the RANK/RANKL/OPG axis^36^, the Fibroblast Growth Factor 23/Klotho axis^37,38^, Tubuloglobulin A^39,40^, osteopontin^41^, and TGF-β^42,43^.

### 3. Shared Genetic Contributors

Genes encoding Matrix Gla Protein have been implicated in the development of both atherosclerosis and OP. Research on mice deficient in the Matrix Gla Protein gene revealed a dual phenotype, characterized by vascular calcifications, reduced bone mineral density (BMD), and an increased incidence of fractures^44^.

### 4. Causal association

Physical activity has also been postulated to be a mediating factor in the relationship between CHD and reduction in BMD. The presence of CHD could restrict levels of physical activity, which in turn may lead to a decrease in BMD.

This investigation acknowledges a number of constraints. Initially, the outcomes from alternative MR methodologies (MR-Egger, weighted median, and mode-based estimations) did not entirely align with those derived from the inverse-variance weighted (IVW) approach in the singular MR analyses. However, in conditions free from heterogeneity and pleiotropy, the IVW estimates are considered the preferred method. Secondly, for the bi-sample MR assessments, the participant pools for the exposure and outcome investigations should be mutually exclusive. The degree of participant overlap in this research was indeterminate. Yet, employing robust analytical instruments (e.g., an F-statistic significantly exceeding 10) can help reduce bias due to sample overlap^45^. Thirdly, while prior MR research indicated a possible causal link of CHD influencing BMD, divergent findings were noted, and the reasons for such discrepancies are yet to be determined. Fourthly, MR analysis segregated by age, sex, and stature was not possible due to the constraints within the GWAS summary statistics. Fifthly, the MR findings were confined to populations of European descent; thus, their extrapolation to Asian ancestries may not be directly transferable.

The prevalence of CHD and OP rises with age, making them leading causes of death and disability. Traditionally, these two conditions have been viewed as unrelated, with their co-occurrence attributed to independent processes associated with aging. However, recent biological and epidemiological evidence suggests a connection between them that goes beyond age and shared risk factors. Some argue that they may share similar molecular, cellular, and biochemical pathways of disease onset. Delving deeper into the link between CHD and OP, and uncovering their mechanisms, may pave the way for novel strategies in treating and preventing OP. From a clinical perspective, subclinical evaluations of OP can be informed by risk stratification for CHD, aiding in the early identification of high-risk OP candidates. Overall, the relationship between CHD and OP remains contentious. Numerous studies provide both supportive and contrary evidence for this connection, which calls for additional research to definitively determine their relationship and the mechanisms underlying it.

## VI. Conclusion

In summary, although previous studies have highlighted various risk factors for OP, including menopause, aging, and hormonal treatments, this article specifically focuses on CHD. However, further research is still required. Recognizing these risk factors is crucial for OP prevention and treatment. Notably, the MR approach is recognized as an effective tool for investigating disease risk factors because it adeptly avoids confounding factors and the bias of reverse causation. The MR approach has identified CHD as a potential risk factor for OP. Further investigation into the underlying mechanisms of this relationship can assist in preventing OP and identifying new therapeutic targets.

## VII. Data availability statement

Datasets accessible to the public are located at the following source: the Integrative Epidemiology Unit (IEU) GWAS database, available online at (https://gwas.mrcieu.ac.uk).

## VIII. Author contributions

The study was conceptualized and designed by CK and HX. ZW, SZ, HG, and GZ were responsible for analyzing the data and drafting the manuscript. Manuscript revisions were undertaken by SY. The collection of data was handled by WW and PX. YC and MY provided the necessary resources. CK led the research team. All authors have made substantial contributions to the work and have given their approval to the final version submitted for publication.

## Data Availability

All data generated or analyzed during this study are included in this published article. Additional datasets used and/or analyzed during the current study are available from the corresponding author on reasonable request

## IX. Acknowledgments

We thank all of the investigators for sharing summary-level data on GWAS. We would like to thank Figdraw (https://www.figdraw.com/) for the drawing help on Figure 1.

